# Variability of mutational signatures is a footprint of carcinogens

**DOI:** 10.1101/2023.11.23.23298821

**Authors:** Maike L. Morrison, Laurane Mangé, Sergey Senkin, Noah A. Rosenberg, Matthieu Foll, Lynnette Fernandez-Cuesta, Nicolas Alcala

**Affiliations:** Department of Biology, Stanford University, Stanford, CA 94305-5020, USA; Rare Cancers Genomics Team, Genomic Epidemiology Branch, International Agency for Research on Cancer/World Health Organization, Lyon, 69372, France; Genomic Epidemiology Branch, International Agency for Research on Cancer/World Health Organization, Lyon, 69372, France

## Abstract

Understanding the genomic impact of carcinogens is fundamental to cancer biology and prevention. However, recent coordinated efforts to detect such fingerprints have been largely unsuccessful, challenging the paradigm that carcinogens induce identifiable mutational signatures. Here we introduce a new method based on statistics from population genetics, signature variability analysis (SVA), that elucidates both the diversity of tumor-causing processes and the heterogeneity of population carcinogen exposure. When we use SVA to re-analyze four prominent studies commonly cited as evidence of nonmutagenic carcinogens, we find that tumors induced by environmental carcinogens do possess mutational signature patterns that distinguish them from spontaneous tumors, even if a specific mutational signature cannot be detected. We find that, across cancers, organs, and model organisms, carcinogen exposure generally increases both the diversity of mutational signatures within a tumor and the homogeneity of signature activity across subjects. Importantly, we show that this increase in signature diversity, far from being a background effect, is associated with the geographic incidence of cancer and can facilitate the acquisition of cancer driver mutations. Our results both encourage a re-examination of the genomic impact of numerous substances and introduce new tools for the analysis of the genomic effects of other substances, potentially influencing carcinogen classifications and cancer prevention policies.

---

The genomic paradigm of carcinogenesis posits that cancer is caused by the sequential acquisition of DNA “driver” alterations in cancer genes, generating progressively more aggressive cell clones that ultimately form a tumor^1–4^. DNA alterations are either due to endogenous processes, such as spontaneous deamination or defective DNA mismatch repair^5–7^, or to exogenous exposure to environmental carcinogens such as UV light or tobacco smoke^8,9^. Under this paradigm, the increased risk of cancer in individuals exposed to environmental carcinogens is due to the direct genotoxic effect of carcinogens, which increase the tumor mutational burden (TMB), thereby increasing the probability of a mutation in a cancer driver gene. For example, the increased risk of cancer due to tobacco smoking is mainly attributed to the effect of smoking on the TMB, with lung adenocarcinoma samples from tobacco smokers having ten-fold the TMB of those from never smokers (∼10 mutations per Mb and∼1 mutation per Mb, respectively^10,11^).

However, several recent studies have found no genomic effect of carcinogen exposure, challenging the ubiquity of the genomic paradigm of carcinognesis. First, exposing mice to 20 suspected or known carcinogens, Riva and colleagues^12^ found that “most of the tested chemicals induced tumors that were indistinguishable from those arising spontaneously”^13^. Second, a large genomic study of lung cancer in never smokers (LCINS) found that patients exposed to second-hand smoke were genomically indistinguishable from unexposed patients, suggesting that second-hand smoke increases cancer risk through nonmutagenic means^11^. Third, our recent analysis of more than 100 malignant pleural mesothelioma tumor whole genome sequences^14^ was the latest in a long line of failures to identify a genomic difference between asbestos-exposed and non-exposed tumors^15,16^, despite asbestos being one of the most powerful known carcinogenic substances^17^. Finally, the most extensive genomic study of cancer across continents to date has suggested that, incidence, like many carcinogen exposures, cannot be explained by genomic differences between tumors^18^. We would naively expect carcinogen exposure to drive both mutational signature activity and cancer incidence. Under this hypothesis, tumors in high-incidence countries should be genomically distinguishable from tumors in low-incidence countries, perhaps having a higher total mutational burden or possessing a unique mutational signature that reflects an exogenous exposure. However, analyzing esophageal cancers across countries with strikingly different incidences, the authors identified no “known or unknown [mutagenic] process that could be responsible for these differences.” Collectively, these studies have caused a shift in focus away from genetic causes of cancer, suggesting that many carcinogens do not directly cause mutations^13^.

At the core of these discoveries stand *mutational signatures*, patterns of mutations throughout the genome that correspond to distinct mutational processes^19–22^. A mutational signature describes the genome-wide relative frequencies of different “mutational types” caused by an exogenous exposure or endogenous process. For example, exposure to tobacco smoke is associated with single-base substitution signature number four (SBS4), a genome-wide pattern of single-base substitutions characterized by high frequencies of C to A substitutions, and low frequencies of other single-base substitution types. In addition to singlebase substitutions (SBS), mutational signatures can be defined in terms of other classes of mutations, such as double-base nucleotide substitutions (DBS signatures), insertions and deletions (ID signatures), copy number alterations (CN signatures), or structural variations (SV signatures). Within a class (e.g., SBS), each “mutational type” corresponds to both a possible alteration (e.g., C to T substitution) and the alteration’s genomic context (e.g., the flanking 5’ and 3’ bases for SBS; see methods for details); the number of mutational types in each signature class typically ranges from 32 for SV signatures to 96 for SBS signatures, and up to 1,536 types in extended contexts.

Mutational signatures are ideally detected through wholegenome sequencing of tumors, whose genomes often contain signatures of multiple mutational processes. For example, in Figure 1, Sample 2 of Population 1 is dominated by SBS4 (Figure 1c), and the tumor’s mutational spectrum is correspondingly also dominated by C to A substitutions (Figure 1a). Analysis of mutational signatures usually begins with the *de novo* extraction of signatures based on the mutational spectra of samples in a cohort and the decomposition of these *de novo* signatures into novel signatures and known signatures based on whether they are present in established databases (e.g., the Catalog Of Somatic Mutations In Cancer, COSMIC)^23^. Subsequent analyses typically focus on the activity of mutational signatures in each sample, also called signature activity. When analyses consider many signatures at once, the signature activities within each tumor sample are usually averaged at the population level for straightforward comparison of signature activity between populations (Figure 1e). Alternatively, each signature may be analyzed in isolation, through the computation of associations between signature presence or population-level mean signature activity, and variables such as cancer incidence or carcinogen exposure^18^.

**Figure 1.**
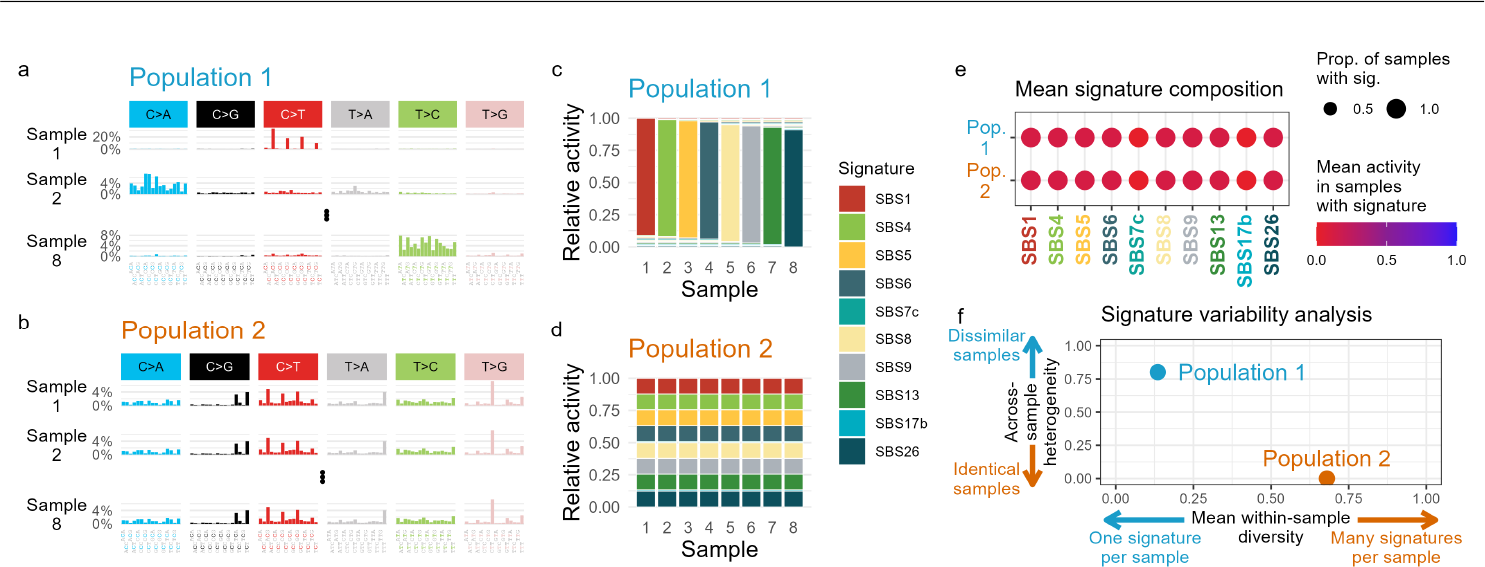
Mutational composition of two hypothetical populations of tumor samples with identical mean signature activities but extremely different variability within and across tumor samples. **a**, Example mutational spectra of hypothetical samples from Population 1. Each sample in Population 1 is dominated by a single mutational signature with just a 1% activity from the remaining nine signatures. **b**, Example mutational spectra of hypothetical samples from Population 2. All samples are caused by an identical mixture of ten mutational signatures. **c-d**, Relative activity of each mutational signature in each sample in Population 1 (c) and Population 2 (d). **e**, Populations 1 and 2 are identical both in their mean relative signature activities and in the proportion of samples with each signature. Dot size corresponds to the proportion of samples in each population that contain a given signature, while color corresponds to the mean relative activity of each signature across all samples in the population. **f**, Populations 1 and 2 have very different values of within-sample diversity (mean expected heterozygosity; computed from equation 6) and across-sample heterogeneity (*F*_*ST*_ ; computed from equation 9).

In this study, we propose a new statistical framework for the analysis of mutational signature data. Signature variability analysis (SVA) quantifies both the diversity of mutational signatures active within each tumor, and the heterogeneity of signature activities across tumors. These two statistics capture patterns of signature activity that are missed when each signature’s activity is analysed in isolation, enabling the detection of carcinogens that affect the diversity of the entire mutational repertoire of a tumor but do not leave a specific mutational signature. In our analyses, SVA reveals previously overlooked genomic impacts of carcinogen exposures and predicts previously unexplained geographic gradients of cancer incidence. We further show how signature diversity can influence carcinogenesis by influencing the occurrence of driver mutations.

## Signature variability analysis reveals patterns obscured by analysis of mean signature activity

To understand signature variability analysis, consider two hypothetical populations, each containing eight tumor samples. In Population 1 each tumor is dominated by a single signature (Figure 1a,c). In Population 2, each tumor contains an identical combination of ten signatures (Figure 1b,d). Tumors in Population 1 were each caused by a distinct large-effect process, while tumors in Population 2 encountered identical exposures to many mutational processes. When these populations are summarized in terms of mean signature activities, as is standard practice^20^ (see signal database^26^), the opposing etiologies driving carcinogenesis are indistinguishable (Figure 1e). However, the differences in population-level mutational signature activity are revealed by considering the within-sample diversity and across-sample heterogeneity of mutational signatures, a framework we call signature variability analysis (SVA, Figure 1f).

SVA draws on a long history of population-genetic statistics devised to understand the genetic diversity within populations and the genetic differentiation across populations^27–30^. When considered jointly, these two dimensions of variability can reveal the patterns of migration, selection, and demographic change experienced by a set of populations^31–33^. Here, we extend these population-genetic statistics to mutational signature data through an analogy between the diversity of alleles within or across populations, and the diversity of mutational signature activities within or across tumors. SVA uses standard measures of within- and across-population genetic diversity: expected heterozygosity and *FST* ^27,28^. We use the expected heterozygosity to measure the diversity of signatures *within* tumors. It is small when each tumor is dominated by a single signature (Population 1) and it approaches 1 as the number of signatures present in each tumor increases, and their frequencies become more even (Population 2, *x*-axis of Figure 1f). We use *F*_*ST*_ to quantify the heterogeneity of mutational signature activity *across* tumors. *F*_*ST*_ is large when each tumor is dominated by a single signature and there are multiple signatures present across tumors (Population 1), and *F*_*ST*_ equals 0 when all tumors have identical signature activities (Population 2, *y*-axis of Figure 1f).

The expected heterozygosity and *F*_*ST*_, which we respectively refer to as within-sample diversity and across-sample heterogeneity, delimit a two-dimensional plane of variability^32^ (Figure 1f). The position of a population of tumors in this space represents both the diversity of mutational processes driving carcinogenesis in the population (within-sample diversity, *x*-axis), and the heterogeneity of population exposure (across-sample heterogeneity, *y*-axis).

We implement SVA in an R package, *sigvar*, which enables the straightforward computation of both within-sample diversity and across-sample heterogeneity, as well as data visualization and statistical hypothesis testing. SVA also allows for the optional inclusion of a pairwise similarity matrix between the signatures, which allows the statistics to account for the varying similarities between pairs of mutational signatures. When not stated otherwise, all analyses presented here account for the similarity (cosine) between mutational signatures^34–36^. The *sigvar* R package is available for download at https://github.com/MaikeMorrison/sigvar.

## Carcinogen exposure increases diversity of mutational signatures

To explore the impact of carcinogen exposure on signature variability, we re-analyzed data from four studies: one in mice and three in humans (Figure 2). First, we re-analyzed data from mice exposed to 20 known or suspected carcinogens, where the resulting tumors were sequenced in order to identify the mutagenic impacts of each substance^12^. Their initial comparison of mean signature activity within carcinogen groups found that most of the carcinogen-induced tumors were indistinguishable from spontaneously occurring tumors (Figure S1a-b). On the contrary, SVA found that all but one carcinogen led to greater within-sample signature diversity than spontaneous tumors, and despite small sample sizes, 10 of the 20 comparisons between carcinogen-induced and spontaneous tumors were statistically significant (Figure 2a; Table S1; bootstrap test with 1,000 replicates, one-sided *P<*0.05 for 10/20 chemicals in liver tumors, and for for 2/9 chemicals in liver tumors). In contrast, only 1 of the 20 carcinogens induced tumors with significantly less within-sample signature diversity than spontaneous tumors (one-sided bootstrap test *P<*0.001). The greater signature diversity in carcinogen-induced tumors was not due to the greater contribution of one signature but rather to the more frequent cooccurrence of many endogenous signatures (e.g., mSBS19 with mSBS1, mSBS5, mSBS40, mSBS12, mSBS17, and mSBS18; Figures S2 and S3, Table S3). SVA also identified a carcinogen that induced liver tumors with more across-sample signature heterogeneity than spontaneous liver tumors (Table S1, one-sided *P<*0.05 for 1 chemical).

**Figure 2.**
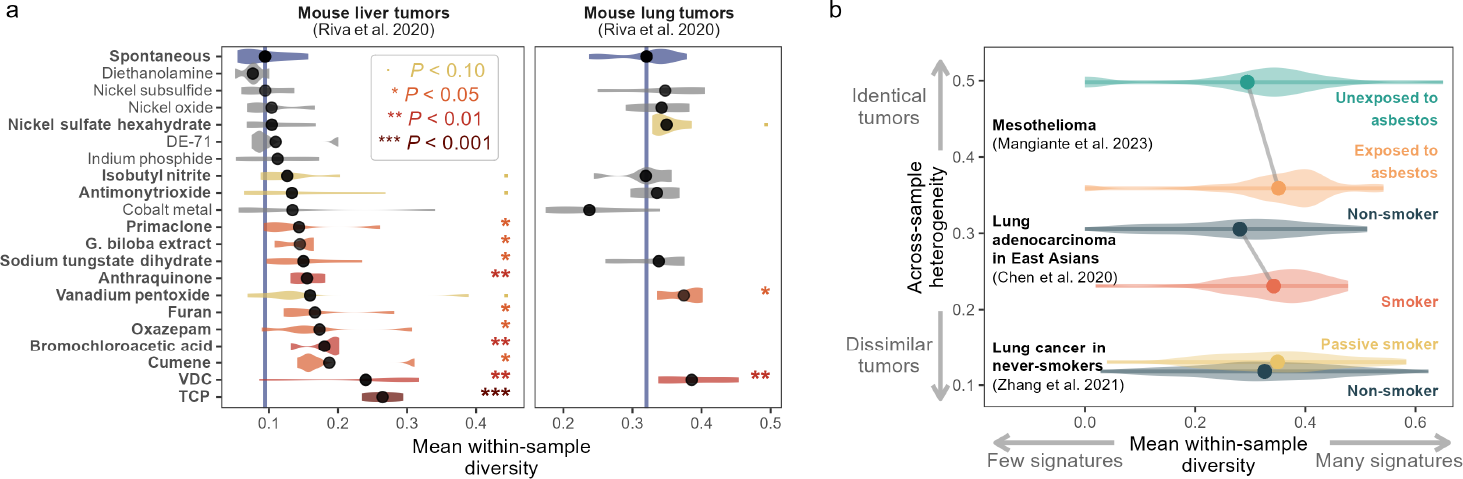
Carcinogen exposure is associated with increased diversity of signature activity within tumors. **a**, Within-sample signature diversity of tumors in mice exposed to 20 known or suspected carcinogenic substances^12^. Violin plots give the distributions of withinsample diversity for all tumors in each exposure group (*n*=3−12 samples per group; Table S2). Vertical lines indicate the mean withinsample diversity in spontaneous tumors for each organ: liver (left) or lung (right). Dots represent the mean within-sample signature diversity for each exposure group. Stars and colors indicate the one-sided significance of bootstrap tests comparing the mean within-sample signature diversity in each exposure group to that of spontaneous tumors. All *P*-values are presented in Table S1. **b**, SVA of tumors in human populations with varying carcinogen exposure. Dots represent the mean within-sample diversity (*x*-axis) and the across-sample heterogeneity (*y*-axis) of signature activities of highand low-carcinogen-exposure populations for three cancer groups: malignant pleural mesothelioma^14^ (orange and green correspond to populations professionally exposed and unexposed to asbestos, *n*=80 and 30, respectively), lung adenocarcinoma in East Asians^24^ (red and navy correspond to smokers and non-smokers, *n*=35 and 53), and lung cancer in never smokers^11^ (yellow and navy correspond to never smokers exposed to second-hand smoke—”passive smokers”—and unexposed never smokers, *n*=64 and 149). Violin plots give the distributions of within-sample diversity (*x*-axis) for all samples from each population.

We next reanalyzed malignant pleural mesothelioma (MPM) whole-genome sequencing data from our recent MESOMICS project^14^. This project sought to identify genomic differences between MPM tumors with varying exposure to asbestos, the primary cause of MPM^17^. The MESOMICS project found that MPM somatic alterations consisted mostly of copy number (CN) alterations, leading us to hypothesize that asbestos exposure would leave a footprint on CN signatures. However, the study failed to identify any difference in mean CN or SBS signature activity between lowand high-exposure patients (Figure S1c-d). Here, we reanalyzed the CN signature data with signature variability analysis. SVA revealed that tumors from patients exposed to asbestos have more within-sample signature diversity and less across-sample heterogeneity than those from unexposed patients (Figure 2b, bootstrap test with 1,000 replicates, onesided *P*=0.028 for within-sample diversity, one-sided *P*=0.022 for across-sample heterogeneity; Figure S4a, S5). This finding posits a previously unknown genomic footprint of asbestos exposure.

Third, we compared the signature variability of LCINS who were exposed to second-hand smoke (“passive smokers”) to those in unexposed never smokers^11^. This study found strong similarities in the mutational patterns between these two exposure groups (Figure S1g-h). While SVA failed to find a significant difference in across-sample signature heterogeneity (bootstrap test with 1,000 replicates, two-sided *P*=0.637), it did find that passive smokers had slightly more diverse mutational signatures than unexposed never smokers (Figure 2b, bootstrap test with 1,000 replicates, one-sided *P*=0.116; Figure S4c, S6). This result is consistent with our findings in mesothelioma and mouse tumors that carcinogen exposure increases signature diversity.

Finally, we reanalyzed data from a study of lung adenocarcinoma in East Asians^24^. As expected, this study found significant differences between the mutational signature activities of smokers and non-smokers, evidence of the well-established direct mutagenic effect of tobacco smoke (Figure S1e-f). Applying SVA to this data, we found that patients who smoke had significantly more within-sample signature diversity and less across-sample heterogeneity than non-smoking patients, suggesting that the causes of adenocarcinoma are at once more diverse within tumors and more homogeneous across tumors from smokers than from non-smokers (Figure 2b, bootstrap test with 1,000 replicates, one-sided *P*=0.112 for across-sample heterogeneity, *P*=0.005 for within-sample diversity; Figure S4b, S7). Interestingly, this finding closely matches the results of SVA between mesothelioma samples from patients exposed and unexposed to asbestos (Figure 2b), despite the fact that the mesothelioma samples do not differ in their mean signature activity (Figure S1c-d). These parallel results suggest previously unappreciated similarities in the genomic effects of carcinogens that do and do not produce a direct mutational signature.

We also investigated whether other clinical characteristics had an effect on signature variability (Table S4). Although patient age is known to influence the TMB^37^, within-sample diversity was not statistically associated with age in any of the datasets (t-tests, *P >* 0.05). This suggests that somatic mutations accumulate with age through the same mutational processes rather than through more and more diverse processes. Mutational processes are not expected to differ by sex, but professional asbestos exposure and smoking are both male-biased. Thus, as expected, males had a significantly greater signature diversity in mesothelioma and lung adenocarcinoma (t-test, *P<*0.05) but not in LCINS. Tumor stage did not have a consistent effect: diversity was similar across stages in lung adenocarcinoma in East Asians, but greater in stages II and III in LCINS (t-test, *P<*0.05). The LCINS cohort was the only dataset including different histological types (adenocarcinoma and carcinoids).The more aggressive adenocarcinoma had a significantly higher diversity than carcinoids (t-test, *P<*2.2 × 10^−16^).

To better understand the effect of carcinogen exposure on signature variability, we simulated signature activities for populations with varying carcinogen exposure (Figure S8). We used the smoking signature activity distributions of the lung adenocarcinoma in East Asians data to derive 10 carcinogen exposure levels ranging from the low exposure of non-smokers to the high exposure of smokers^24^. Our simulations corroborate the results of mesothelioma and lung adenocarcinoma (Figure 2b), demonstrating that within-sample diversity increases and across-sample heterogeneity decreases as carcinogen exposure strengthens (Figure S8a). The one exception to this pattern occurs for very high exposures, when the within-sample diversity reaches a maximum and begins to decrease as one signature (SBS4) starts to strongly dominate all the others (Figure S8b).

In summary, in both *in vivo* murine models and human populations, highly exposed populations consistently had more within-sample diversity than their less exposed counterparts. More highly exposed human populations additionally often had less across-sample heterogeneity.

## Mutational signature variability explains geographic patterns of cancer incidence

In the previous section, we found that populations exposed to carcinogens generally had higher within-sample signature diversity and lower across-sample signature heterogeneity than unexposed populations. In this section, we apply SVA to data from populations with varying cancer incidence. If signature variability influences carcinogenesis, as suggested by the results of the previous section, we would expect high- and low-incidence countries to follow the same SVA patterns as highand lowcarcinogen-exposure populations. We test this hypothesis using data from the Mutographs project^18,25^. This consortium has recently sought to identify mutational signature differences driving global distributions of esophageal squamous cell carcinoma (ESCC) and clear cell renal cell carcinoma (ccRCC), whose dramatically varying geographic incidences are not fully explained by known risk factors.

The study of 552 ESCC samples found similar mean signature profiles across all eight countries, concluding that, if there is an exogenous mutagen driving differences in ESCC incidence, it left no genetic signature^18^ (Figure 3a). However, SVA computed at once across SBS, DBS, and ID signatures reveals a dramatic difference in the signature variability of high- and low-incidence countries. ESCC cases in high-incidence regions from five countries (as defined in the original studies, incidence rates above 20 in 100,000) are each caused by a more diverse set of mutational signatures than those in low-incidence countries (incidence rates below 6 in 100,000), and these signatures are also more homogeneous across tumors (Figure 3b-c, Figure S11; Welch’s two Sample t-test, one-sided *P*=0.001 for mean within-sample diversity, one-sided *P*=0.004 for across-sample heterogeneity; Table S5).

**Figure 3.**
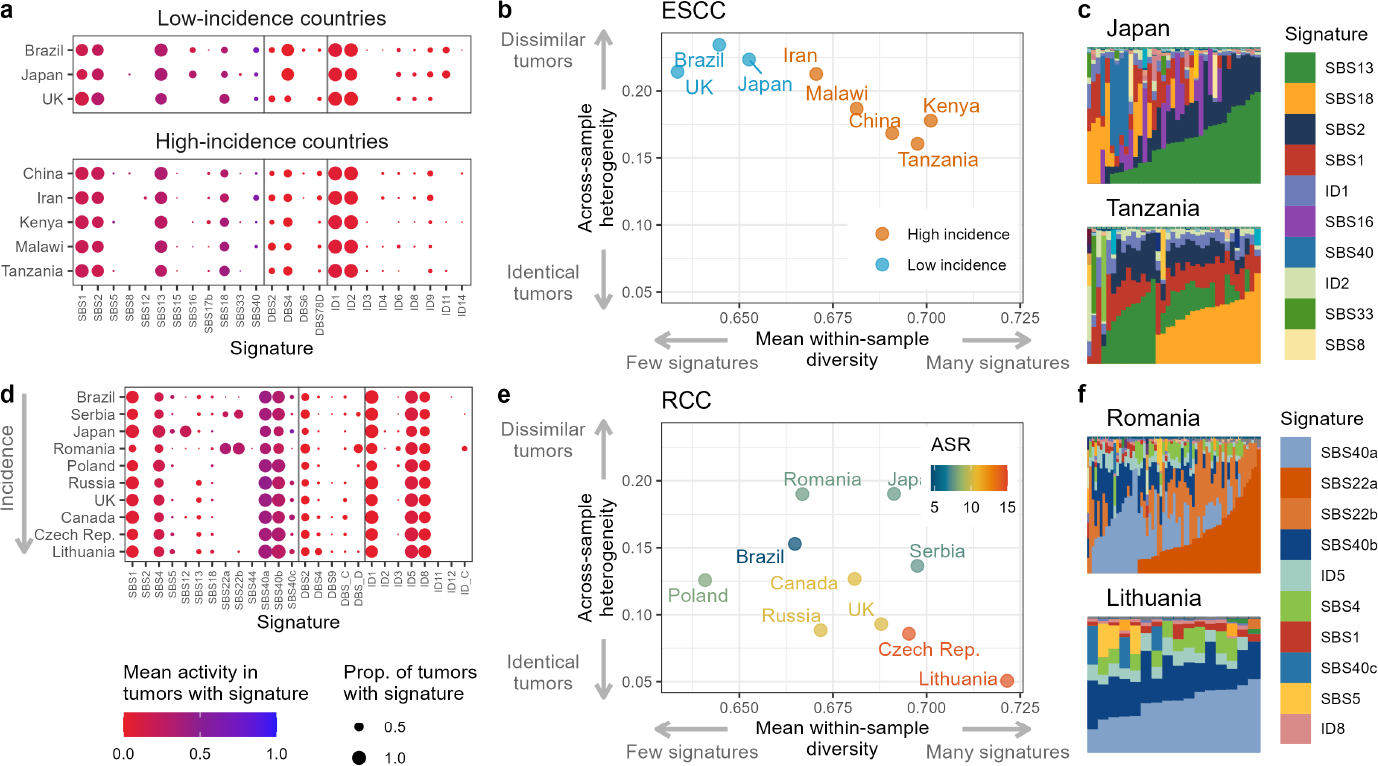
Across cancers and countries, within-sample signature diversity and across-sample signature heterogeneity are associated with incidence. **a**, Mean signature activities across esophageal squamous-cell carcinoma (ESCC) samples from countries with high or low ESCC incidence (*n*=552^18^). **b**, SVA of ESCC samples grouped by country. Colors represent cancer incidence. **c**, Mutational signature activities for samples from Japan and Tanzania. The legend presents the 10 most abundant signatures. **d**, Mean signature activities across clear cell renal cell carcinoma (ccRCC) samples from countries listed in order of increasing ccRCC incidence (*n*=957^25^). **e**, SVA for ccRCC samples grouped by country. Colors represent the age-standardized rate (ASR) of ccRCC in each country. **f**, Mutational signature activities for samples from Romania and Lithuania. Note that in panels b and e, SVA does not account for cosine similarity because the ccRCC results included SBS signatures defined in terms of both 3and 5-nucleotide contexts, complicating the computation of cosine similarities between signatures (see Figure S9 for similar results collapsing all signatures to a 3-nucleotide context and weighting by cosine similarity among signatures). See Figure S10 for a representation of the distributions of within-sample diversity for each country.

The study of 962 clear cell renal cell carcinomas (ccRCC) from 11 countries did find evidence of multiple exogenous exposures^25^, including signatures of aristolochic acid (SBS22a and b) in Romania, Serbia, and Thailand, a signature of unknown origin in Japan (SBS12), and a signature of unknown origin associated with incidence (SBS40b)^25^. However, the mean activities of many signatures were still quite comparable across populations (Figure 3d). Applying SVA to ccRCC, we find that tumors from higher-incidence countries have more within-sample diversity and less across-sample heterogeneity (Figure 3e-f, Pearson correlation, one-sided *P*=0.033 for within-sample diversity, onesided *P*=0.003 for across-sample heterogeneity). This result corroborates our analysis of ESCC (Figure 3b), again finding a positive relationship between within-sample diversity and incidence, and a negative relationship between across-sample heterogeneity and incidence.

In both ESCC and RCC, we find that, across countries, the mean activities of several mutational signatures are associated with SVA results (Tables S6 and S7). For example, we found that the mean activity of SBS40b, which was shown in the original study of RCC to correlate positively with cancer incidence^25^, was significantly negatively correlated with across-sample heterogeneity (Pearson correlation, *R*=−0.708, one-sided *P*=0.011). This result is consistent with our hypothesis that signature composition is more homogeneous in higher incidence countries.

In summary, we find a nearly identical pattern when applying SVA to global samples of ESCC and ccRCC: in both cancer types, higher incidence countries have greater mean within-sample signature diversity and lesser across-sample signature heterogeneity (Figure 3b,e). This result suggests that mutational processes are both more diverse and more ubiquitous in high-incidence countries. In addition, the patterns of signature variability between high- and low-incidence countries parallel those between populations exposed to high and low levels of asbestos or tobacco smoke (Figure 2b). This similarity supports the hypothesis that the genomes of cancers in both high-carcinogen-exposure and high-cancer-incidence populations are directly influenced by carcinogenic exposures, both in cases when an exposure leaves a detectable signature (e.g., smoking, ccRCC) and in cases when no unique signature is detected (e.g., asbestos exposure, ESCC). This symmetry provides further evidence for a link between carcinogen exposure, signature diversity, and carcinogenesis.

## Mutational signature diversity impacts driver mutations

The results presented so far have established that, in nearly all cases, carcinogen exposure and cancer incidence are positively associated with the within-sample diversity of mutational signatures. We hypothesize that this association is mechanistically driven by a relationship between the diversity of a tumor’s mutational signature activity—which determines the total spectrum of mutations acting on the genome—and the probability that a mutation from this spectrum is capable of initiating carcinogenesis—that is, that a mutation is a so-called driver mutation^38^. The greater the overlap between a tumor’s mutational spectrum (e.g., Figure 4a-b) and the spectrum of driver mutations in a cancer-associated gene (e.g., Figure 4c-d; data from the Intogen database^39^), the higher the probability that a mutation in that tumor is a driver mutation in the specified gene.

**Figure 4.**
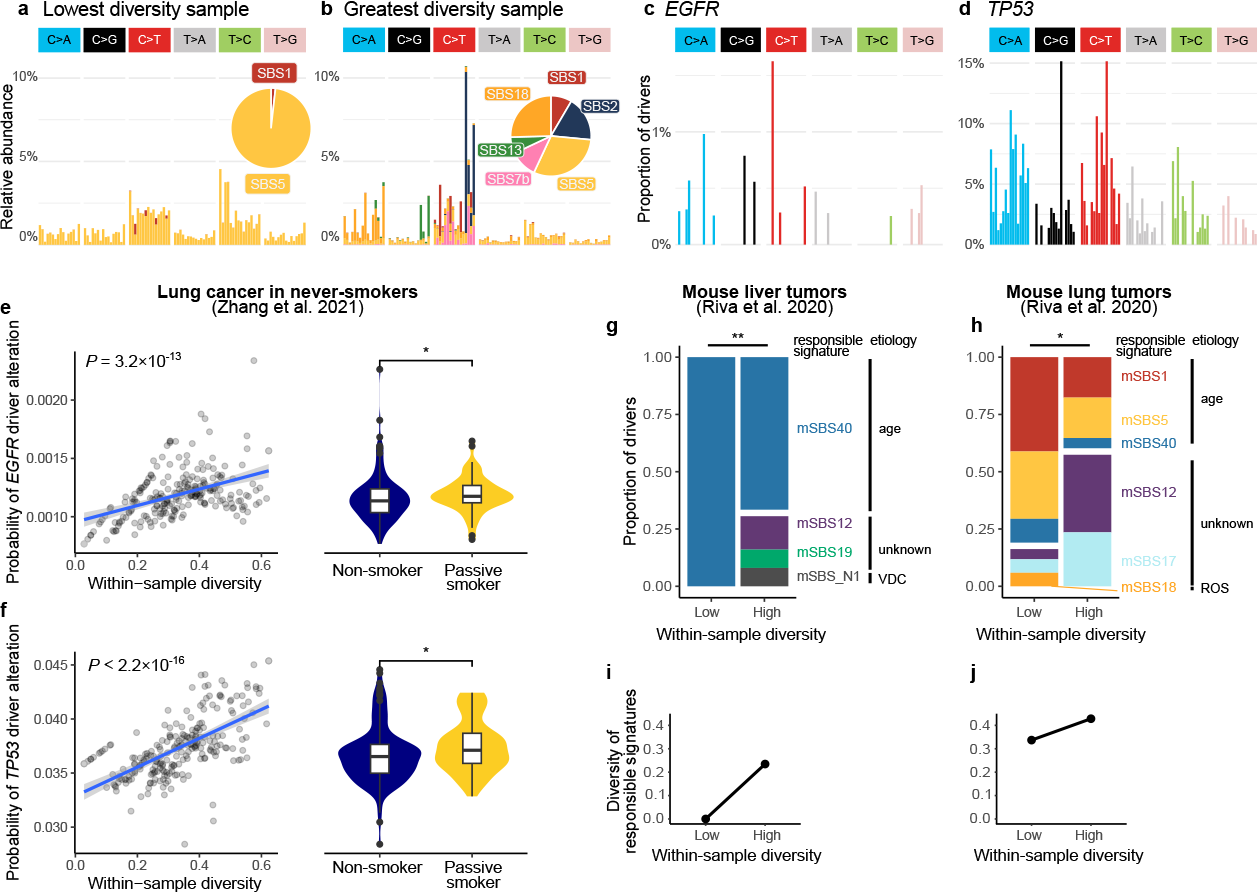
Within-sample diversity impacts driver mutations. **a-b**, SBS mutational spectra of the sample from LCINS^11^ with the least (a) and greatest (b) mutational signature diversity, colored by activity of SBS mutational signatures. **c**-**d**, SBS mutational spectrum of known driver alterations in genes *EGFR* (*n*=25 mutations) and *TP53* (*n*=202 mutations), respectively. **e**, Left: association between within-sample diversity and the probability of a mutation being a driver mutation in gene *EGFR* in LCINS (*n*=232 patients). The *P* value corresponds to a two-sided Spearman correlation test. Right: association between passive smoking and the probability of a driver alteration in *EGFR*; stars indicate the significance level of two-sided Wilcoxon rank sum tests: * for *P<*0.05, * * for *P<*0.01, and* * * for *P<*0.001. In box plots, the center line represents the median, box limits represent the upper and lower quartiles, whiskers represent 1.5 times the interquartile range, and points represent outliers. **f**, Same as **e** but for gene *TP53*. **g**, Mean activity of signatures most likely to be responsible for the recurrent driver mutations observed in mouse liver tumors^12^, across two groups of samples: samples with below-median within-sample diversity (Low, *n*=24 mutations) and samples with above-median diversity (High, *n*=25 mutations). **h**, Same as **g** but for lung tumors (Low, *n*=17 mutations; High, *n*=17). ROS: reactive oxygen species. In g-h, a white horizontal bar separates age-associated and non-age-associated signatures, and stars correspond to Fisher’s exact tests of the proportions of age-associated vs non-age-associated tumors. **i**-**j**, Within-sample diversity of the mean signature activities in driver mutations presented in g-h.

To test whether signature diversity is associated with driver mutation probability, we compute the driver mutation probability for LCINS samples^11^ in each of the two most frequently mutated genes, *EGFR* and *TP53* (Table S8). We found that, across these tumor samples, the driver mutation probability was indeed positively correlated with the within-sample diversity of mutational signatures (*P*=3.2×10^−13^ for *EGFR* and *P<*2.2^−16^ and for *TP53*; Table S9). Mutations in tumors with a diversity of 0.6 had a 1.15 to 1.20 times greater probability of being drivers compared to tumors with a diversity of 0.1 (Fig. 4e-f). Interestingly, passive smokers, adenocarcinomas, and samples from the molecular clusters with the greatest number of copy number alterations (Forte and Mezzo-forte^11^) had the highest probability of driver mutations in both *EGFR* and *TP53* (two-sided Wilcoxon rank sum tests *P<*0.05, Figure 4e-f and Figure S12a-d). Comparing the mutational spectrum of the lowest-diversity sample (Fig. 4a) to that of the greatest-diversity sample (Fig. 4b), we see that the increased driver mutation probability in the greatest-diversity sample is mostly due to the combined influence of two mutational processes. On the one hand, mutations attributed to APOBEC enzymes (SBS13, dark blue bars in Fig. 4b) created an increase of C to T mutations that could hit many known driver positions (red bars in Fig. 4c-d). On the other hand, mutations attributed to reactive oxygen species (SBS18, dark yellow bars in Fig. 4b) created an excess of C to A mutations that could hit another set of known drivers (blue bars in the right panels in Fig. 4c-d).

We independently replicated the correlation between the probability of driver mutations and within-sample signature diversity using the mice experiments from Riva et al.^12^ (Table S10), and the ESCC tumors from Moody et al.^18^ (Tables S11 and S12). We show that a majority of the main drivers found in liver and lung cancers of mice exposed to carcinogens (*Hras, Braf, Fgfr2*) are more likely to occur in diverse tumors (Figure S12e-g). Interestingly, drivers in *Kras* in lung tumors are negatively correlated with within-sample diversity; this opposite effect of diversity on *Kras* and *Fgfr2* drivers is coherent with the observation from Riva and colleagues that *Kras* and *Fgfr2* mutations are mutually exclusive. Similarly, a majority of drivers from ESCC were more likely to occur in diverse tumors (*PIK3CA, KMT2D, NFE2L2, EP300*; Figure S13c-e, g), and some were only more likely to occur in diverse tumors in low-incidence countries (*TP53, CDKN2A*; S13a,b). This effect of diversity on driver occurrence probability resulted in a significant difference in driver mutation probability between tumors from lowand high-incidence countries (Figure S13). Note that all these results consider the probability that a single alteration hits a driver mutation, so it imposes a similar TMB to all samples; additionally, in most of these datasets, no significant association between carcinogen exposure or incidence and TMB were reported (Figure S14, Table S13). Together, our results exemplify the potential for diverse mutational repertoires (e.g., comprising both SBS13 and 18) to together increase the driver mutation probability, thus heightening cancer risk even without increasing mutational burden.

To further verify that signature diversity indeed impacts driver mutations, we turned to the data of Riva and colleagues^12^, who determined the mutational signature most likely to be responsible for each driver mutation identified in the sequenced tumors. Grouping tumors into above- or below-median signature diversity groups, we analyzed the proportion of drivers caused by each mutational signature (Figure 4g-h). We found that, for both liver and lung tumors, driver mutations in the highsignature-diversity group were caused by a more diverse array of responsible signatures (Figure 4g-j). For example, drivers in the low-diversity liver cancer group (left bar in Figure 4g) are all due to mSBS40, so the diversity of the group is 0 (left dot in Figure 4i). Drivers in the high-diversity group, on the other hand, are due to multiple signatures, causing their diversity to be greater (right dot in Figure 4i). We also found that the abundances of signature etiologies differed significantly between high- and low-diversity tumors (Fisher’s exact test, *P*=0.004 and 0.032 for liver and lung, respectively). In liver cancers, while all 24 drivers identified in low-diversity tumors were generated by a single signature—age-associated signature mSBS40—the 25 drivers found in high-diversity tumors were generated by four different signatures, including endogenous signatures of unknown etiology (mSBS12, mSBS19) and a signature associated with VDC exposure (mSBS_N1; Figure 4g). Similarly, in lung cancers, drivers from low-diversity samples are predominantly generated by age-related signatures (mSBS1, mSBS5, and mSBS40), while drivers in high-diversity samples are mostly generated by signatures of unknown etiology (mSBS12 and mSBS17; Figure 4h).

### Discussion

In this study, we introduce signature variability analysis (SVA), a new statistical framework for the analysis of mutational signature activity in a population. Using this framework, we show that most carcinogens do have a direct effect on DNA, but that, rather than producing a unique signature that increases the TMB, they influence multiple mutational processes simultaneously, increasing the *diversity* of signatures observed in a tumor, and in many cases increasing the homogeneity of populationwide signature activities as well. Through analyses of data from murine carcinogen exposure experiments^12^ and human populations exposed to asbsestos or tobacco smoke^11,14,24^, we find that populations with higher carcinogen exposure tend to have more diverse mutational signature activities, regardless of whether a unique signature was associated with the exposure. We also find that, in human populations, smokers and people exposed to asbestos have more homogeneous signature activities than do non-smokers and unexposed people. Using data from the Mutographs project^18,25^, we also show that differences in geographic prevalence of ESCC and ccRCC are both associated with higher within-sample diversity and lower across-sample heterogeneity of mutational signatures. The symmetry in the effects of carcinogen exposure and cancer incidence on mutational signature variability provides evidence for a link between exposure, signature variability, and carcinogenesis. Indeed, we use known driver alterations to show that the diversity of mutational processes can influence the probability that a tumor-initiating mutation occurs.

Our results suggest a novel conceptual mode of action for many carcinogens. Most of the mutational signatures that we report as affected by carcinogens are associated with endogenous processes, such as APOBEC activity (SBS2 and 13) and DNA repair (SBS3). This relationship suggests that carcinogens may disrupt endogenous cellular processes. While the stimulation of endogenous processes by carcinogens has been suggested as a possible explanation for their lack of an exogenous signature^13^, SVA allows for a precise quantification of this effect. We find that strong exposures (e.g., tobacco smoke^24^, asbestos^14^, murine carcinogen experiments^12^) lead to stronger signals of mutational signature variability than weaker exposures such as secondhand smoke[11]. For the latter, we find that the genomic effect of second-hand smoke can only be revealed by an in-depth analysis of the impact of diversity on driver mutations in genes *EGFR* and *TP53*.

Our results have profound implications for cancer prevention. By enabling the discovery of elusive footprints of exposure, SVA opens the door to discovering more associations between environmental exposure and DNA damage. Genotoxicity, DNA repair alteration, and the induction of genomic instability are considered key characteristics of carcinogens^40,41^. However, evidence for activity of these phenomena is usually limited to descriptions of specific signatures. Our results suggest that these phenomena could also be defined by the diversity of the mutational spectrum.As an example, we report that bromochloroacetic acid, currently classified as only “possibly carcinogenic to humans” (group 2B), increases the diversity of mutational signatures more than most of the 20 other substances tested by Riva and colleagues^12^, including three group 1 carcinogens^42^. Furthermore, all three substances tested by Riva and colleagues^12^ that were mutagenic based on Ames tests but were not associated with a mutational signature or an increase in TMB (isobutyl nitrate, G. biloba extract and primaclone) led to elevated signature diversities, possibly reconciling genomic analyses with classical genotoxicity tests. In fact, most substances increased diversity relative to spontaneous tumors in at least one tissue.

Our results do not preclude the possibility that many carcinogens also act by non-mutagenic means. For example, cancer driver mutations have been found in healthy tissue^43,44^, suggesting that these mutations do not cause cancer in isolation and prompting researchers to envisage an alternative model of carcinogenesis in which a carcinogen acts primarily by altering the tumor micro-environment rather than by causing direct DNA damage^13,45^. This so-called promoter model of carcinogenesis is reminiscent of the evolutionary model of adaptation from the standing genetic variation^46,47^. Under both models, the “initiator” of natural selection (mutations) and the “promoter” (environmental change) are provided by independent processes. The cancer promoter model was recently empirically supported by the discovery that air pollution promotes lung cancer in never smokers^48^ and that chronic inflammation promotes leukemia^49^. Our discovery of increased signature diversity in response to carcinogen exposure suggests that carcinogens could simultaneously act as initiators, by increasing the diversity of the mutational repertoire and thus the probability of driver mutations, and as promoters by modifying the tumor microenvironment.

Our results were enabled by adopting the emerging paradigm that the biology of cancer can be viewed from the perspective of population biology, a field in which variability at all levels— among cells, individuals, or groups of individuals—is treated as a central component of a system. Population biology perspectives that consider variability within and among tumors have contributed to many recent discoveries in cancer biology. For example, viewing a tumor as a population of cancer cells has revealed the importance of early events in the aggressiveness and spatial organization of some tumors^50^and the potential to harness tumor-cell ecological interactions to optimize treatment^51^. Embracing the diversity of molecular profiles in cancer patient populations is a core principle of personalized medicine, guiding cancer treatment approaches such as neoantigen-based therapeutic cancer vaccines^52^ and individual-based prevention models^53^. Our study similarly embraces a population-level view to place variability—of somatic alterations within an individual tumor and of mutational signature activities across tumors—as main objects of interest. Indeed, we found that carcinogens can give rise to population-level phenomena in the variability of signature activity even when they do not lead to detectable individual-level properties, such as presence of an exogenous signature in a given sample. Given the morphological and molecular heterogeneity observed within and across tumors^54^ and the heterogeneity among cancer patients in prognosis and treatment response^55^, our study supports the role of population biology perspectives as key to the future of cancer research.

## Methods

### Mutational data

We retrieved publicly available mutational signature activities and definitions from cancer whole-genome sequencing studies. We focused on the most common types of somatic alterations affecting cancer genomes^21^: single base substitutions (SBS), double-base substitutions (DBS), indels (ID), and copy number alterations (CN). We here briefly describe these types of data; see for example^56^ for a more detailed description of mutational data. For each type of somatic alteration, alterations are classified into a number *X* of channels or types; a set of alterations is then represented as a vector of size *X* called the “mutational spectrum,” with positive integer entries corresponding to the number of alterations of each type. In this paper, we focus our analysis on a normalized mutational spectrum, with entries between 0 and 1 corresponding to the relative abundance of alterations of each type.

### Single base substitutions

Single base substitutions (SBS) are single-nucleotide mutations in which a DNA base-pair is replaced by another base-pair. For example, an A:T base-pair can be substituted by a C:G base-pair; following standard practice, we use the notation A:T>C:G, simplified to T>G using the pyrimidine of the Watson-Crick pairs as reference. With this notation, there are 6 possible substitutions (C>A, C>G, C>T, T>A, T>C, and T>G). Following standard practice^21^, we use an extended classification taking into account the 5’ and 3’ flanking nucleotides, called the “three-nucleotide context.” Under this classification, each possible substitution (e.g., C>A) has sixteen possible contexts (ACA>AAA, ACC>AAC, ACG>AAG, ACT>AAT, CCA>CAA, CCC>CAC, CCG>CAG, CCT>CAT, CCA>CAA, GCC>GAC, GCG>AAG, TCT>AAT, TCA>AAA, TCC>AAC, TCG>AAG, and TCT>TAT). This leads to a mutational spectrum with *X* = 6 × 16 = 96 channels.

### Double base substitutions

Double base substitutions (DBS) are simultaneous alterations of two adjacent nucleotides. An example DBS is AA:TT>GG:CC, which is simplified to TT>CC using the standard notation which lists the pair of bases with the most pyrimidine nucleotides. Listing all source and substituted strand-agnostic doublets, resolving palindromes and cases with equal numbers of pyrimidine nucleotides, leads to the standard *X* = 78 channels used in most studies.

### Indels

Indels (ID) comprise small insertions and deletions of one or more nucleotides (usually fewer than 100). For example, C:G>-:-, simplified to C>-, for a single nucleotide deletion. Indels are usually classified based on the size of the indel (1, 2, 3, 4, 5, and 6 or more) and the type of region where it occurred. Regions considered are repetitive regions (regions with a similar sequence repeated multiple times), and microhomology regions (regions with a DNA sequence partially matching that of the focal deletion); note that these two types of regions are usually used for classification of indels because of their connection to mutational processes important to cancer such as DNA repair deficiency and microsatellite instability^20^. We thus used the usual *X* = 83 channels, which account for indel type, length, and region type.

### Copy number alterations

Copy number (CN) alterations correspond to large (usually more than 100bp) duplications or deletions of chromosomal segments. For example, the loss of a segment of 10kb in chromosome 3, leading to a region with only a single copy (complete loss of the paternal or maternal copy), or the gain of 5 copies of a segment of 1kb in chromosome 5, leading to a region with a copy number of 6 (1 copy of the maternal or paternal chromosome, and 5 copies of the other chromosome). CN were recently classified using the size of the segment (0–100kb, 100kb–1Mb, 1Mb–10Mb, 10Mb–40Mb, greater than 40Mb), its number of copies (0, 1, 2, 3-4, 5-8, 9 and more), and its heterozygosity (heterozygous if at least one copy of the paternal and maternal chromosome are present, and loss of heterozygosity otherwise)^22^. This leads to *X* = 48 channels.

### Mutational spectrum and signatures

The full set of mutations in a tumor or patient sample can be summarized in terms of the abundance of each SBS, DBS, ID, or CN channel, synthesizing genome-wide data into a set of categories that consider the alteration and its immediate context, but ignore each mutation’s absolute position in the genome. For example, because every single-base substitution in the genome belongs to one of the 96 SBS channels, the genome-wide SBS data can be summarized by the abundance of each of the 96 SBS types. This list of 96 type abundances is known as the tumor or sample’s SBS “mutational spectrum.” Mutational spectra can be generated for DBS, ID, and CN alterations as well. Figure 1a-b give examples of SBS mutational spectra.

Mutational signatures, genome-wide patterns of mutations often associated with a particular mutational process, are also defined in terms of these SBS, DBS, ID, or CN types. For example, the SBS mutational signature associated with tobacco smoking, SBS4, has high abundances of C to A substitutions and low abundances of other substitution types. Mutational signatures are defined through both large-scale population sequencing and controlled exposure experiments in order to both identify the signatures of population-level mutational processes and match these signatures to causes^20,57^.

The mutational spectrum of a tumor or patient sample can be represented as a weighted sum of mutational signatures. The weight of each signature, known as the signature’s “activity,” represents how many mutations in the tumor or patient sample are attributed to each signature. For example, a tumor might contain 500 mutations from SBS5 and 500 mutations from SBS40, in which case the tumor mutational spectrum would be approximately equal to 500 times the number of mutations in each SBS channel in the SBS5 spectrum, plus 500 times that in the SBS40 spectrum. In this paper, we focus our analysis on the “relative activity” of each signature, which is the activity of each signature normalized by the total mutational burden: in the previous example, the relative activity would be 0.5 for SBS5 and 0.5 for SBS40. In general, we use “activity” to refer to “relative activity” in this paper. In standard inferences of mutational signatures, the mutational spectrum of each individual is approximated as a combination of established mutational signatures, such as those in the catalog of somatic mutations in cancer (COSMIC^58^) database, which contains the mutational signatures known to be frequently present in cancer samples, referred to as the COSMIC signatures. There are about 100 SBS COSMIC signatures, about 10 DBS COSMIC signatures, and about 20 ID or CN COSMIC signatures.

### Cosine similarity between mutational signatures

It is often of interest to quantify the similarity between mutational signatures, for example to check whether signatures identified in different populations are the result of the same mutational process, or to account for the challenge of confidently distinguishing between similar signatures^59^. The cosine similarity is the standard metric to quantify the similarity between mutational signatures^23^; it corresponds to the angle *θ* between two vectors **x** and **y** of length *n*, computed as the dot product of the vectors divided by the product of the norm of the two vectors ||**x**|| and ||**y**||^60^:

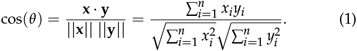

Because in our case **x** and **y** represent mutational signatures with non-negative values, their dot product and thus the cosine similarity range in [0, 1].

## Datasets

We used the SBS (96 types) and CN (48 types) COSMIC signatures called in 115 malignant pleural mesothelioma samples from the French MESOBANK (Supplementary Table from^14^) and corresponding signature definitions (COSMIC3.1, Table S35 from^14^). We used the 8 SBS COSMIC signatures and 3 de novo signatures from mice exposed to 20 different carcinogens (github repository https://github.com/team113sanger/mouse-mutatation-signatures containing the data and scripts from^12^), focusing on the 58 lung and 112 liver tumors because they were the only ones including spontaneous tumors that serve as a control. We used the 14 SBS signatures from 232 LCINS, among which 213 have passive smoking information (personal communication with authors from^11^ to obtain the source data for Figure 4 panel 6) and corresponding signature definitions (COSMIC v3, Table S7 from^11^); note that in the original paper and thus here, samples where SBS4 was detected were removed from the analysis. We used the SBS signatures from *n*=88 lung adenocarcinoma in East Asians from the CHG cohort^24^. We used the COSMIC SBS, DBS, and ID signatures from 552 ESCC samples from 8 countries (Table S15 from^18^) and the SBS, DBS, and ID signatures from 957 out of the 962 clear cell RCC samples from 11 countries (Table S6 from^25^; we excluded samples from Thailand because of their small sample size of *n* = 5). The RCC signatures include 3-nucleotide-context COSMIC signatures as well as the 5-nucleotide-context, de novo SBS signatures that we were not well described by the existing COSMIC catalog. All mutational signatures were measured from distinct samples.

## Signature variability analysis

### Computing variability summary statistics

The statistics that form the foundation of signature variability analysis are standard, widely used measures in population genetics: heterozygosity and *F*_*ST*_^28,61^. In standard applications, these two statistics are used to quantify the within-population diversity or across-population heterogeneity of allele frequencies—a scenario mathematically analogous to the relative activities of signatures within or across tumor samples. Let *q*_*i,k*_ represent the relative activity of signature *k* in sample *i*, **q**_**i**_ = (*q*_*i*,1_, *q*_*i*,2_, …, *q*_*i,K*_) be the signature profile of sample *i*, and **Q** be a matrix with rows **q**_**1**_, **q**_**2**_, …, **q**_**I**_ representing the relative signature activity of all *I* samples in the population. Note that the relative activity of each signature is between 0 and 1 (0 ≤ *q*_*i,k*_ ≤ 1) and the relative activities of all signatures for each sample must sum to 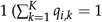 for each *i*). We define within-sample signature diversity using a statistic known as the expected heterozygosity^61^,

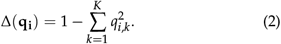

This statistic quantifies the diversity of all *K* signatures in a single tumor sample, **q**_**i**_. Δ(**q**_**i**_) = 0 when **q**_**i**_ contains a single signature with activity 1 and all others have activity 0 (approximately each sample in Figure 1c). Δ(**q**_**i**_) →1 as the number of signatures increases and their activities get more even (see each sample in Figure 1d). Taking the average over all *I* tumor samples in the population of tumor samples, **Q**, gives the mean within-sample diversity, the *x*-axis of Figure 1f,

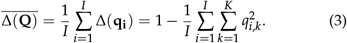

We quantify across-sample heterogeneity with an extension of the population-genetic statistic *F*_*ST*_^61^. This statistic is traditionally used to quantify variability in allele frequencies across populations, but has been recently used to quantify variability in other contexts, such as ancestry inference^36^. This statistic is defined in terms of both 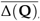, the mean within-sample diversity, and 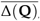, the variability of the pooled signature activities across all samples. The variability of the pooled signature activities is computed by first computing the average activity of each signature across all samples, and then computing the diversity of this pooled activity vector:

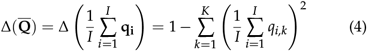

*F*_*ST*_ is then defined as the normalized difference between this pooled diversity and the mean within-sample diversity^62^,

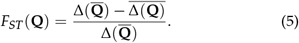

*F*_*ST*_ = 1 when each sample contains a single signature, since each individual has no diversity and thus 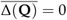 (approx. Figure 1c). *F*_*ST*_ = 0 when all samples are identical in their signature activities, so the mean within-sample diversity, 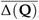, is identical to the variability of the pooled signature activities, 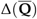 (Figure 1d).

We use a previously developed extension of biological diversity statistics^35^ to allow these two statistics, 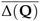 and *F*_*ST*_, to account for the cosine similarity between mutational signatures. Let **S** be a *K*×*K* matrix with entries *s*_*k,l*_ equal to the cosine similarity between signature *k* and signature *l*. **S** is symmetric (*s*_*k,l*_ = *s*_*l,k*_) and its diagonal elements equal 1, since each signature is identical to itself (*s*_*k,k*_ = 1). Every entry *s*_*k,l*_ is between 0 and 1, equalling 0 when signatures *k* and *l* are totally dissimilar and equalling 1 when they are identical. We incorporate this similarity matrix into our previous definition for the expected heterozygosity:

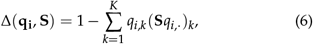

where *q*_*i*,·_ is the vector (*q*_*i*,1_, …, *q*_*i,K*_) and 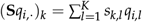 represents the weighted average similarity between signature *k* and a random signature in sample *i. q*_*i,k*_ ≤ (**S***q*_*i*, ·_)_*k*_ ≤1, with (**S***q*_*i*, ·_)_*k*_ equalling *q*_*i,k*_ when signature *k* is totally dissimilar from every other signature, and equalling 1 when signature *k* is identical to every signature. This extension is identical to that used by^35^, who incorporated a similarity matrix into a diversity statistic very similar to this one in order to develop a biodiversity measure that accounted for species similarity. Incorporating this signature-similarity-aware definition of heterozygosity into 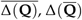, and *F*_*ST*_ (**Q**), we have:

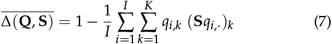

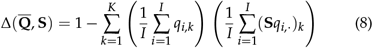

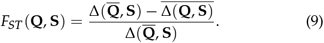

When **S** = **I**_*K*×*K*_, the *K*-dimensional identity matrix which has all *K* diagonal elements equal to 1 and all off-diagonal elements equal to 0, every signature is treated as identical to itself and totally dissimilar from other signatures, and hence 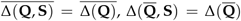, and *F*_*ST*_ (**Q, S**) = *F*_*ST*_ (**Q**). In this paper, we use a similarity matrix **S** whose entries are defined as the cosine similarity between all pairs of mutational signatures active in a population of tumor samples. However, in principle any symmetric matrix with diagonal elements equal to 1 and off-diagonal elements between 0 and 1 could be used (e.g., the L2 norm), giving the user flexibility in their signature definitions and weightings.

### Bootstrap protocol for comparing variability statistics

Bootstrapping is a standard statistical procedure that involves sampling with replacement from a data set in order to generate a null distribution against which one can test hypotheses^63^. We use bootstrapping to test whether two populations of tumor samples, **Q**_**1**_ with *I*_1_ samples, and **Q**_**2**_ with *I*_2_ samples, have significantly different mean within-sample signature diversity or across-sample signature heterogeneity (Figure 1f). Our null hypothesis is that there is no difference in diversity or heterogeneity between **Q**_**1**_ and **Q**_**2**_. We use bootstrapping to test this hypothesis through the following steps:

1. **Merge the two matrices to generate a null distribution of tumor samples**. Call this matrix **Q**_null_, with *I*_1_ rows equal to **Q**_**1**_ and *I*_2_ rows equal to **Q**_**2**_.
2. **Draw “bootstrap replicate matrices” from the null distribution**. Randomly draw *I*_1_ rows with replacement from **Q**_null_ to generate a bootstrap replicate of **Q**_**1**_, 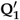, and ran-domly draw *I*_2_ rows with replacement from **Q**_null_ to generate a bootstrap replicate of **Q**_**2**_, **Q**^′^_**2**_. Because each of these replicate matrices is drawn from **Q**_null_, there should be no true difference in their mean within-sample diversity or across-sample heterogeneity.
3. **Compute the mean within-sample diversity or acrosssample heterogeneity for** 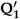 **and** 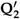. Then compute the difference in mean within-sample signature diversity 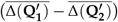 and the difference in across-sample heterogeneity 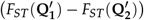 between the two populations.
4. **Repeat steps 2-3 many times** (typically between 100 and 1,000 replicates). This procedure generates a bootstrap distribution of the difference in mean within-sample diversity and a bootstrap distribution of the difference in *FST* under the null hypothesis that there is no true difference between **Q**_**1**_ and **Q**_**2**_. Example bootstrap distributions for the data in Figure 2b are presented in Figure S4.
5. **Compute a** *P***-value for each statistic by comparing the observed difference between the two matrices to the bootstrap distribution of the difference**. The one-way *P*-value is computed as the proportion of bootstrap replicate differences in the statistic that are either less than or greater than the observed difference. The two-way *P*-value is the proportion of bootstrap replicate differences whose absolute value is greater than the absolute value of the observed difference.

Note that such a bootstrapping protocol is the standard way of comparing *F*_*ST*_ and other diversity statistics in population genetics (see recommendations in^29^, and popular softwares *fstat*^64^, *hierfstat*^65^, and *genepop*^66,67^). See Figures S15 and S16 for an example showing that the sample size does not influence the results of the test, and Figures S17 and S18 for both empirical and simulated datasets showing that the test leads to uniform *P* value distributions.

### The *sigvar* R package

The signature variability analysis R package, *sigvar*, contains functions to compute within-sample signature diversity and across-sample signature heterogeneity (function *sigvar*) and to conduct hypothesis testing via bootstrapping (function *sigboot*). The package also contains data visualization functions that can reproduce all figures in this manuscript. These functions can visualize signature proportions across samples (*plot_sig_prop*; Fig. 1c,d, Fig. 3c,f), SBS mutational spectra (*plot_SBS_spectrum*, Fig. 1a,b, Fig. 4a-d), dot-plots of the mean activities of signatures across a set of samples (function *plot_dots*, Fig. 3b,e), and bootstrap distributions of differences in signature variability between different groups (function *sigboot*, Fig. S4). We have also written vignettes providing tutorials to reproduce all results and figures from this study (see function *vignette(‘sigvar’)*). The package will be submitted to bioconductor shortly.

## Statistical tests

Statistical tests were performed using R v4.1.2. Differences between mean values of a categorical variable were tested using t-tests when data was approximately normally distributed, or Wilcoxon rank-sum tests otherwise. The association between two continuous variables was tested using Pearson correlation if the residuals were approximately normally distributed, and using Spearman correlations otherwise; whenever clinical covariables (e.g., sex, age) were available, we fit multivariable models in addition to univariable linear models. Test statistics, effect sizes, degrees of freedom and *P* value of tests are reported in supplementary tables whenever appropriate.

### Investigating the effect of purity on SVA

For the datasets where sample purity data was available, we checked whether purity was influencing our conclusions using Pearson correlation tests. For the ESCC data, purity was defined using the allele-specific copy number analysis of tumors (ASCAT) algorithm, as presented in the original study^18^.

#### Lung cancer in never smokers

We show in Figure S19a-b that purity does not drive differences in SVA results between nonsmokers and passive smokers. When each sample is analyzed individually, sample purity is slightly negatively associated with within-sample diversity (Figure S19a). This association is not significant among passive smokers (Pearson correlation, *R*= − 0.14, *P*=0.27) but is significant among non-smokers (Pearson correlation, *R*=−0.24, *P*=0.0028). Nevertheless, there is no significant difference in purity between passive smokers and non-smokers (Welch two-sample two-sided t-test, *P*=0.42; Figure S19b).

#### Mesothelioma as a function of asbestos exposure

We similarly show in Figure S19c-d that purity does not drive differences in SVA results between mesothelioma tumors from exposed and unexposed individuals. When each sample is analyzed individually, sample purity is slightly negatively associated with within-sample diversity among exposed and unexposed individuals (Figure S19c) but these associations are not significant (Pearson correlations *P >* 0.60). Additionally, there is no significant difference in purity between exposed and unexposed individuals (Welch two-sample two-sided t-test, *P*=0.62; Figure S19d).

#### ESCC across countries with varying cancer incidence

We show in Figure S20 that purity does not drive differences in SVA results between high- and low-ESCC-incidence countries. Consistent with the main results of Figure 3, when each sample is analyzed individually (instead of being aggregated by country), there is a significant difference in within-sample diversity between all samples in high-incidence countries and all samples in low-incidence countries (Welch two-sample two-sided t-test, *P*=0.016; Figure S20a). However, there is no significant difference in purity between all samples in high-incidence countries and those in low-incidence countries (Welch two-sample twosided t-test, *P*=0.881; Figure S20b), suggesting that the difference in diversity is not driven by a difference in purity. When each sample is analyzed individually, sample purity is significantly positively associated with within-sample diversity (Figure S20c). This association is stronger in high-incidence countries (Pearson correlation, *R*=0.14, *P*=0.002) than in low-incidence countries (Pearson correlation, *R*=0.18, *P*=0.146). However, this association does not hold when samples are aggregated by country and the mean purity and diversity are analyzed. There is no significant relationship between the mean purity of samples in a country, and the mean within-sample signature diversity of those samples (Pearson correlation, *R*=−0.26, *P*=0.534; Figure S20d). There is also no significant relationship between the mean purity of samples in a country, and the across-sample signature heterogeneity of those samples (Pearson correlation, *R*= − 0.06, *P*=0.885; Figure S20e).

### Simulations

#### SVA as a function of exposure intensity

In order to understand the effect of carcinogen exposure on mutational signature variability, we constructed a simple model of signature activity parameterized by the lung adenocarcinoma data from an East Asian population of both smokers and non-smokers^24^. This study includes the activities of three signatures: aging, APOBEC, and smoking.

First, we fit a beta distribution to the distribution of each signature’s activity in the study population. The aging signature was distributed Beta(*α*=1.06, *β*=88.35) (*SE*_*α*_ =0.14, *SE*_*β*_ =14.86), yielding a mean activity of 0.012. The APOBEC signature was distributed Beta(*α*=0.27, *β*=35.94) (*SE*_*α*_ =0.03, *SE*_*β*_ =8.45), yielding a mean activity of 0.007. Because smokers and non-smokers had very different smoking signature activity distributions, we fit two separate beta distributions for the smoking signature. For smokers, this distribution was Beta(*α*=0.47, *β*=14.66) (*SE*_*α*_ =0.09, *SE*_*β*_ =4.51), with a mean activity of 0.031. For non-smokers, this distribution was Beta(*α*=0.21, *β*=34.02) (*SE*_*α*_ =0.03, *SE*_*β*_ =11.21), with mean 0.006.

Second, in order to consider intermediate smoking exposure, we interpolated eight sets of beta distribution parameters (*α, β*) at even intervals between the parameters of the smokers’ and the non-smokers’ smoking signature activity distributions. This procedure resulted in ten beta distributions for smoking signature activity, ranging from strong exposure (smokers, high mean activity) to low exposure (non-smokers, low mean activity).

Third, we used these beta distributions to simulate the signature activity of lung adenocarcinoma samples with varying smoking exposure. For each sample, we independently drew one exposure from the aging distribution, one from the APOBEC distribution, and one from one of the smoking distributions. The choice of which smoking distribution to draw from was determined by the desired exposure strength for the sample. We repeated this procedure in order to simulate populations of many samples. For each smoking exposure level, we simulated 500 populations, each with 500 samples. In total, this involved simulating 2,500,000 samples: 10 smoking exposure levels 500×populations 500×samples per population.

Finally, we used the sigvar R package to compute the across-sample and mean within-sample diversity of the normalized signature activities in each simulated population (Figure S8). We find that across-sample heterogeneity decreases monotonically with exposure strength. Mean within-sample diversity, however, increases until an intermediate threshold exposure strength just less than the exposure of a smoker, after which it begins to decrease. This peak in diversity happens when the mean normalized activity of each sample’s most abundant signature is minimized.

#### SVA in negative control simulations

In order to show that SVA did not detect spurious signature variability when none was present, we simulated data from two groups of samples drawn from the same underlying population, with signature activities parameterized by the LCINS data^11^.

First, we fit a Dirichlet distribution to the signatures of never smokers unexposed to second-hand smoke (149 samples from^11^). To do so, we fit the mean vector *α* to the observed mean signature attributions. This vector was then given as an input to the method “rDirichlet.acomp” (from R package compositions^68^) to simulate 213 samples, just as in the original dataset. Second, for each of the 100 repetitions, the 213 simulated samples were shuffled and separated into two subsets. The two subsets simulated and compared contain the same number of samples as the non-smokers (149 samples) vs passive smokers (64 samples) from the data. We finally used the sigvar R package to compute within- and across-sample variabilities for each repetition and compute the *P* values of the difference between the two groups (Figure S18). We find that *P* value distributions are uniform, as expected under the null hypothesis.

### Computing the probability of driver mutations

We compute the probability that a random single base substitution creates a known driver alteration using a random mutation model conditional on the driver gene mutational spectrum and the tumor sample’s mutational spectrum.

#### Notation

This analysis relies on comparing two sets of mutational spectra: the mutational spectrum occurring in tumor samples (see e.g., Figure 4a-b) and the spectrum of driver mutations in established cancer genes (see e.g., Figure 4c-d). We represent the mutational spectrum of tumor sample *i* by a vector with dimensions 1×96, **m** = (*m*_ACA*>*AAA_, *m*_ACC*>*AAC_, …, *m*_TTT*>*TGT_), where for example *m*_ACA*>*AAA_ corresponds to the proportion of all single-base substitutions in the tumor sample that are ACA>AAA. All entries *m*_XYZ*>*XWZ_, where X, Y, and Z are nucleotides, are positive and sum to 1. We denote by **d** = (*d*_ACA*>*AAA_, *d*_ACC*>*AAC_, …, *d*_TTT*>*TGT_) the 1×96 vector of the proportion of driver mutations of each type, with 0 ≤ *d*_*XYZ>XWZ*_ ≤1 the proportion of known *Y > W* drivers with context *XYZ* divided by the total number of positions with context *XYZ, n*_*XYZ*_, in the focal cancer gene.

#### Mutational model

We consider a mutational model where a single mutation is drawn at random from a given mutational spectrum **m**, using a multinomial model. Under this model, the probability Pr(*XYZ > XWZ*) that a mutation *XYZ*>*XWZ* (e.g., ACA>AAA) is one of the 96 SBS types is equal to the proportion of *XYZ*>*XWZ* mutations in the spectrum, *m*_XYZ*>*XWZ_. We also assume that mutations are spatially uniform, so a mutation of type *XYZ*>*XWZ* is equally likely to hit any genomic position with a *Y* nucleotide in a *XYZ* context. We further assume that among all possible genomic positions, a known number *J* can lead to driver alterations if substituted by the right nucleotide.

#### Probabilities

We first seek to determine the probability of occurrence of a given driver mutation *Y > W* at a specific genomic position *j* in a focal gene, within a *X*_*Z* context (*W, X, Y*, and *Z* being nucleotides), and given a tumor sample’s mutational profile **m**. We denote this probability by Pr(*XY*_*j*_ *Z > XW*_*j*_ *Z*|**m**). We compute this probability as the product of the probability for a mutation to be of type *XYZ* given the sample’s mutational spectrum **m**, Pr(*XYZ>XWZ*|**m**), and the probability for a mutation of type *XYZ > XWZ* to happen at genomic position *j*, Pr(*XY*_*j*_ *Z > XW*_*j*_ *Z* |*XYZ > XWZ*).

As detailed above, under our mutational model, Pr(*XY*_*j*_ *Z > XW*_*j*_ *Z*|**m**) is simply the proportion of *XYZ*>*XWZ* mutations in the sample’s mutational spectrum, *m*_XYZ*>*XWZ_. For example, in the mutational spectrum of the lung cancer sample presented in Figure 4a, TCT>TTT substitutions only account for 2% of substitutions; thus, the probability that a random mutation drawn from this spectrum is of type TCT>TTT is 2%. On the contrary, TCT>TTT substitutions account for 7% of substitutions in the sample presented in Figure 4b, so the probability for a mutation to be of type TCT>TTT is 7% for this sample.

Under the spatially uniform mutation model, the probability for a mutation of type *XYZ*>*XWZ* to happen at a given genomic position *j*, Pr(*XY*_*j*_ *Z > XW*_*j*_ *Z*|*XYZ > XWZ*), is equal to 1/*n*_*XYZ*_, where *n*_*XYZ*_ is the number of positions with context *XYZ* (e.g., ACA) in the focal gene. For example, the C>T driver mutation in chromosome 7 at position 55,171,191 (cancer gene *EGFR*) is of the type TCT>TTT (rightmost red bar in Figure 4c). Because there are around 200 positions in *EGFR* with such TCT contexts, the probability that a random TCT>TTT mutation hits position 55,171,191 is approximately 0.5%.

As a result, given a mutational spectrum **m**, the probability that an *XYZ*>*XWZ* mutation occurs at location *j* is thus

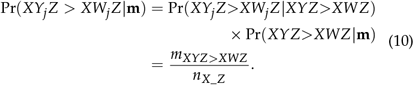

We now consider the set of all *J* driver alterations of a focal gene, denoted by *XY*_*j*_ *Z*>*XW*_*j*_ *Z* with *j* ranging from 1 to *J*.

Summing Pr(*XY*_*j*_ *Z > XW*_*j*_ *Z*|**m**) from eq. 10 across all *J* driver alterations, and using vector notation, we obtain the probability that a random mutation is a driver alteration:

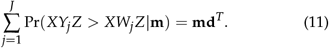

#### Data application

##### Lung cancer in never smokers

To illustrate the impact of signature diversity on the probability of a driver mutation in LCINS, we focused on genes *EGFR* and *TP53*. We computed **m** from observed profiles from^11^, by summing the mutational profiles of COSMIC signatures weighted by their activity **q**_*i*_ in each sample. We computed **d** using the list of known driver alterations from the Intogen database^39^, and computing the number of positions of each context using the sequence of the main transcript of each gene from the Ensembl database (ENST00000275493.7 and ENST00000269305.9, respectively).

##### Mice exposed to known or suspected carcinogens

To illustrate the impact of signature diversity on the probability of a driver mutation in mice exposed to known or suspected carcinogens, we focused on genes reported as recurrently mutated in the study, *Hras* and *Braf* in liver tumors, and *Fgfr2* and *Kras* in lung tumors. We computed **m** from observed profiles from^12^, by summing the mutational profiles of COSMIC signatures weighted by their activity **q**_*i*_ in each sample. We computed **d** using the list of driver alterations from the paper (Figure 4), and computing the number of positions of each context using the sequence of the main transcript of each gene from the Ensembl database (ENSMUST00000026572.11 for *Hras*, ENSMUST00000111710.8 for *Kras*, ENSMUST00000122054.8 for *Fgfr2*, and ENSMUST00000002487.15 for *Braf*).

##### ESCC accross countries

We focused on the most frequently mutated driver genes identified in the cohort^18^: *TP53, CDKN2A, PIK3CA, KMT2D, NFE2L2, NOTCH1, EP300*. We computed **m** from observed profiles from^18^, by summing the mutational profiles of COSMIC signatures weighted by their activity **q**_*i*_ in each sample. We computed **d** using the list of driver alterations from the paper, and computing the number of positions of each context using the sequence of the main transcript of each gene from the Ensembl database.

## Ethics and inclusion

This study is based on the reanalysis of published data, so we refer readers to the original publications for information about inclusion of researchers from the locations where the research was conducted.

## Reporting summary

Further information on research design is available in the Reporting Summary linked to this article.

## Data Availability

All data are publicly available (see cited references).

## Code Availability

R code to produce the figures is available in the vignettes of package *sigvar* at https://github.com/MaikeMorrison/sigvar.

## Acknowledgements

This work is part of the Rare Cancers Genomics Initiative (www.rarecancersgenomics.com), and results shown are in part based on data generated by the initiative in the context of the MESOMICS project. Financial support was provided by a France-Stanford Center for Interdisciplinary Studies grant (NAR, NA, and MF), a US Department of Defense grant (RA210061 to MF and LFC), a French Agence Nationale de la Recherche grant (Tremplin program 2023 to LFC), a National Science Foundation Graduate Research Fellowship (MLM), the Anne T. and Robert M. Bass Stanford Graduate Fellowship (MLM), and a PhD fellowship from the French Ligue Nationale Contre le Cancer (LM). We are grateful for constructive conversations and comments from many lab mates and colleagues, including Chloe Shiff.

## Authors’ Contributions

NA designed the study. NA, MLM, and LM analysed the data. SS provided advice on mutational signature analyses. MF, NAR, and LF-C provided feedback on the results. NA and MLM wrote the manuscript. All authors reviewed the manuscript.

## Additional information

**Supplementary Information** is available for this paper (see below in combined PDF).

## References

1. Boveri, T. Zur Frage der Entstehung Maligner Tumoren [Origin of malignant tumors]. Gustav Fisher, Jena (1914).

2. Berenblum, I. in Etiology: Chemical and Physical Carcinogenesis (1982).

3. Nowell, P. C. The clonal evolution of tumor cell populations. Science 194 (1976).

4. Knudson, A. G. Genetics of human cancer. Annual review of genetics 20 (1986).

5. Liu, B. et al. Analysis of mismatch repair genes in hereditary non-polyposis colorectal cancer patients. Nature Medicine 2 (1996).

6. Pfeifer, G. P. Mutagenesis at methylated CpG sequences in Current Topics in Microbiology and Immunology 301 (2006).

7. Welch, J. S. et al. The origin and evolution of mutations in acute myeloid leukemia. Cell 150 (2012).

8. Pfeifer, G. P. et al. Tobacco smoke carcinogens, DNA damage and p53 mutations in smoking-associated cancers. Oncogene 21-48 (2002).

9. Pfeifer, G. P., You, Y. H. & Besaratinia, A. Mutations induced by ultraviolet light. Mutation Research - Fundamental and Molecular Mechanisms of Mutagenesis 571 (2005).

10. Alexandrov, L. B. et al. The repertoire of mutational signatures in human cancer. Nature 578 (2020).

11. Zhang, T. et al. Genomic and evolutionary classification of lung cancer in never smokers. Nature Genetics 53, 1348–1359 (2021).

12. Riva, L. et al. The mutational signature profile of known and suspected human carcinogens in mice. Nature Genetics 52, 1189–1197 (2020).

13. Balmain, A. The critical roles of somatic mutations and environmental tumor-promoting agents in cancer risk. Nature Genetics 52, 1139–1143 (2020).

14. Mangiante, L. et al. Multiomic analysis of malignant pleural mesothelioma identifies molecular axes and specialized tumor profiles driving intertumor heterogeneity. Nature Genetics 55, 607–618 (2023).

15. Bueno, R. et al. Comprehensive genomic analysis of malignant pleural mesothelioma identifies recurrent mutations, gene fusions and splicing alterations. Nature Genetics 48 (2016).

16. Creaney, J. et al. Comprehensive genomic and tumour immune profiling reveals potential therapeutic targets in malignant pleural mesothelioma. Genome Medicine 14 (2022).

17. Van Zandwijk, N., Rasko, J. E., George, A. M., Frank, A. L. & Reid, G. The silent malignant mesothelioma epidemic: a call to action. The Lancet Oncology 23 (2022).

18. Moody, S. et al. Mutational signatures in esophageal squamous cell carcinoma from eight countries with varying incidence. Nature Genetics 53, 1553–1563 (2021).

19. Nik-Zainal, S. et al. Mutational processes molding the genomes of 21 breast cancers. Cell 149 (2012).

20. Alexandrov, L. B. et al. The repertoire of mutational signatures in human cancer. Nature 578, 94–101 (2020).

21. Koh, G., Degasperi, A., Zou, X., Momen, S. & Nik-Zainal, S. Mutational signatures: emerging concepts, caveats and clinical applications. Nature Reviews Cancer 21, 619–637 (2021).

22. Steele, C. D. et al. Signatures of copy number alterations in human cancer. Nature 606, 984–991 (2022).

23. Islam, S. M. et al. Uncovering novel mutational signatures by de novo extraction with SigProfilerExtractor. Cell Genomics 2 (2022).

24. Chen, J. et al. Genomic landscape of lung adenocarcinoma in East Asians. Nature Genetics 52, 177–186 (2020).

25. Senkin, S. et al. Geographic variation of mutagenic exposures in kidney cancer genomes. medRxiv 14, 2023.06.20.23291538. https://www.medrxiv.org/content/10.1101/2023.06.20.23291538v1 (June 2023).

26. Degasperi, A. et al. Substitution mutational signatures in whole-genome–sequenced cancers in the UK population. Science 376 (2022).

27. Wright, S. The genetical structure of populations. Annals of Eugenics 15, 323–354 (1951).

28. Nei, M. Analysis of gene diversity in subdivided populations. Proceedings of the National Academy of Sciences of the United States of America 70, 3321–3323 (1973).

29. Weir, B. S. & Cockerham, C. C. Estimating F-Statistics for the Analysis of Population Structure. Evolution 38 (1984).

30. Holsinger, K. E. & Weir, B. S. Genetics in geographically structured populations: Defining, estimating and interpreting FST. Nature Reviews Genetics 10 (2009).

31. Pickrell, J. K. et al. Signals of recent positive selection in a worldwide sample of human populations. Genome Research 19, 826–837 (2009).

32. Wang, J. Does GST underestimate genetic differentiation from marker data? Molecular Ecology 24, 3546–3558 (2015).

33. Matthey-Doret, R. & Whitlock, M. C. Background selection and FST: Consequences for detecting local adaptation. Molecular Ecology 28, 3902–3914 (2019).

34. Cavalli-Sforza, L. L. & Edwards, A. W. F. Phylogenetic analysis: models and estimation procedures. American Journal of Human Genetics 19, 233–257 (1967).

35. Leinster, T. & Cobbold, C. A. Measuring diversity: the importance of species similarity. Ecology 93, 477–489 (2012).

36. Morrison, M. L., Alcala, N. & Rosenberg, N. A. FSTruct: An FST-based tool for measuring ancestry variation in inference of population structure. Molecular Ecology Resources 22, 2614–2626 (2022).

37. Campbell, P. J. et al. Pan-cancer analysis of whole genomes. Nature 578. ISSN: 14764687 (2020).

38. Stratton, M. R. Exploring the genomes of cancer cells: Progress and promise. Science 331 (2011).

39. Martínez-Jiménez, F. et al. A compendium of mutational cancer driver genes. Nature Reviews Cancer 20 (2020).

40. Smith, M. T. et al. Key characteristics of carcinogens as a basis for organizing data on mechanisms of carcinogenesis. Environmental Health Perspectives 124 (2016).

41. International Agency for Research on Cancer. IARC Monographs on the Identification of Carcinogenic Hazards to Humans - Preamble. World Health Organisation (2019).

42. World Health Organization. Some chemicals that cause tumours of the urinary tract in rodents. IARC Monographs on the Evaluation of Carcinogenic Risks to Humans 119 (2019).

43. Moore, L. et al. The mutational landscape of normal human endometrial epithelium. Nature 580 (2020).

44. Martincorena, I. et al. Somatic mutant clones colonize the human esophagus with age. Science 362 (2018).

45. Lopez-Bigas, N. & Gonzalez-Perez, A. Are carcinogens direct mutagens? Nature Genetics 52 (2020).

46. Hermisson, J. & Pennings, P. S. Soft sweeps: Molecular population genetics of adaptation from standing genetic variation. Genetics 169 (2005).

47. Barrett, R. D. H. & Schluter, D. Adaptation from standing genetic variation. Trends in Ecology & Evolution 23, 38–44 (2008).

48. Hill, W. et al. Lung adenocarcinoma promotion by air pollutants. Nature 616, 159–167 (2023).

49. Rodriguez-Meira, A. et al. Single-cell multi-omics identifies chronic inflammation as a driver of TP53-mutant leukemic evolution. Nature Genetics (2023).

50. Sottoriva, A. et al. A big bang model of human colorectal tumor growth. Nature Genetics 47 (2015).

51. West, J. et al. Towards multidrug adaptive therapy. Cancer Research 80 (2020).

52. Blass, E. & Ott, P. A. Advances in the development of personalized neoantigen-based therapeutic cancer vaccines. Nature Reviews Clinical Oncology 18 (2021).

53. Adams, S. J. et al. Lung cancer screening. The Lancet 401, 390–408 (2023).

54. Hoadley, K. A. et al. Cell-of-Origin Patterns Dominate the Molecular Classification of 10,000 Tumors from 33 Types of Cancer. Cell 173 (2018).

55. Bray, F. et al. Global cancer statistics 2018: GLOBOCAN estimates of incidence and mortality worldwide for 36 cancers in 185 countries. CA: A Cancer Journal for Clinicians 68 (2018).

56. Bergstrom, E. N. et al. SigProfilerMatrixGenerator: A tool for visualizing and exploring patterns of small mutational events. BMC Genomics 20 (2019).

57. Alexandrov, L. B., Nik-Zainal, S., Wedge, D. C., Campbell, P. J. & Stratton, M. R. Deciphering Signatures of Mutational Processes Operative in Human Cancer. Cell Reports 3, 246–259 (2013).

58. Sondka, Z. et al. The COSMIC Cancer Gene Census: describing genetic dysfunction across all human cancers. Nature Reviews Cancer 18 (2018).

59. Bergstrom, E. N., Barnes, M., Martincorena, I. & Alexandrov, L. B. Generating realistic null hypothesis of cancer mutational landscapes using SigProfilerSimulator. BMC Bioinformatics 21 (2020).

60. Timm, N. H. Applied Multivariate Analysis (Springer-Verlag, New York, 2002).

61. Gillespie, J. H. Population Genetics: A Concise Guide 2nd ed. (The Johns Hopkins University Press, Baltimore, Maryland, 2004).

62. Alcala, N. & Rosenberg, N. A. Mathematical constraints on FST: Multiallelic markers in arbitrarily many populations. Philosophical Transactions of the Royal Society B 377, 20200414 (2022).

63. Efron, B. & Tibshirani, R. An Introduction to the Bootstrap (Chapman and Hall, New York, 1993).

64. Goudet, J. FSTAT (Version 1.2): A Computer Program to Calculate F-statistics. Journal of Heredity 86 (1995).

65. Goudet, J. HIERFSTAT, a package for R to compute and test hierarchical F-statistics. Molecular Ecology Notes 5 (2005).

66. Raymond, M. & Rousset, F. GENEPOP (Version 1.2): Population Genetics Software for Exact Tests and Ecumenicism. Journal of Heredity 86 (1995).

67. Rousset, F. GENEPOP’007: A complete re-implementation of the GENEPOP software for Windows and Linux. Molecular Ecology Resources 8 (2008).

68. Van den Boogaart, K. G. & Tolosana-Delgado, R. “compositions”: A unified R package to analyze compositional data. Computers and Geosciences 34 (2008).

