## Supplementary for "Variability of mutational signatures is a footprint of carcinogens"

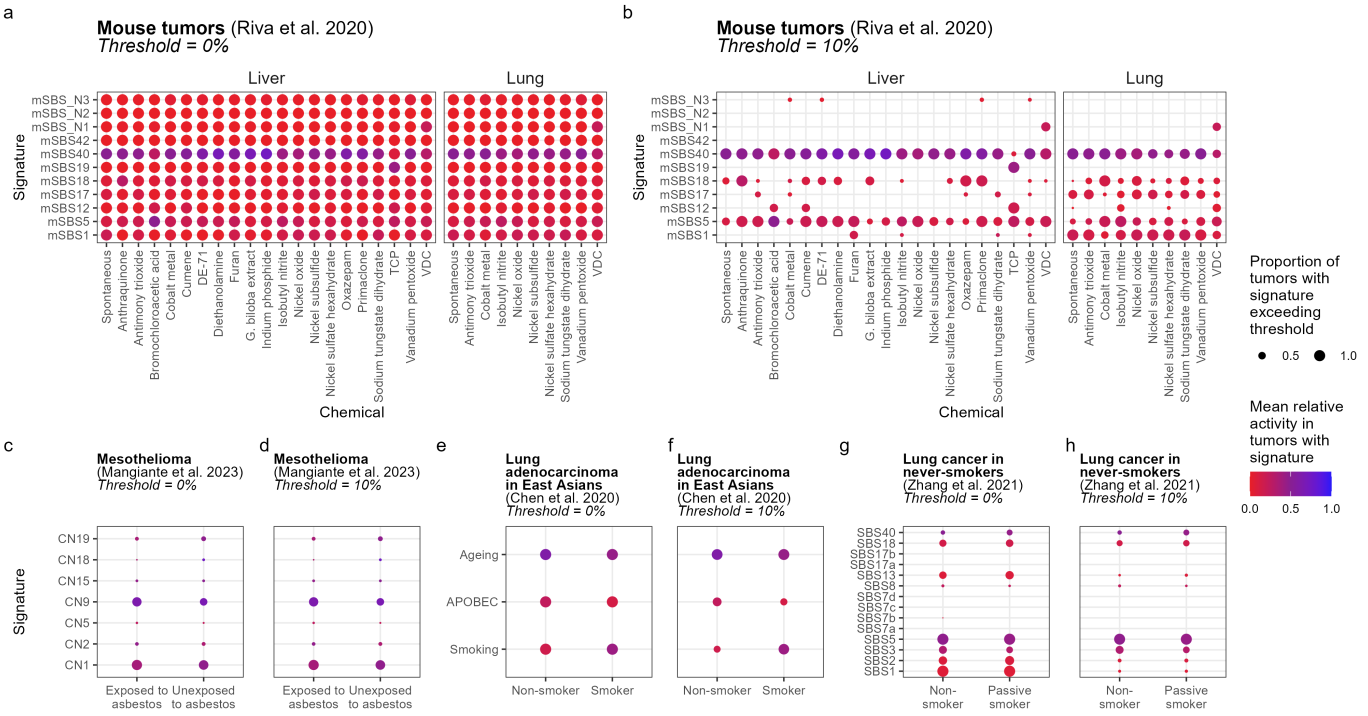


**Figure S1 | Mean signature activity in samples with varying carcinogen exposure.** SVA results of these datasets are presented in Figure 2. **a,** Mean signature activity in liver (left) or lung (right) tumors in mice exposed to 20 suspected or known carcinogens. Data from[12]. **b,** a, but restricted to signatures present at greater than 10% per sample. **c,** Mean signature activity in human mesothelioma samples from populations exposed and unexposed to asbestos. Data from[14]. **d,** c, but restricted to signatures present at greater than 10% per sample. **e,** Mean signature activity in lung cancers of smokers and non-smokers; data from[24]. **f,** e, but restricted to signatures present at greater than 10% per sample. **g,** Mean signature activity in LCINS exposed to second-hand smoke ("passive smoker") and unexposed never smokers ("non-smoker"); data from[11]. **e,** g, but restricted to signatures present at greater than 10% per sample.

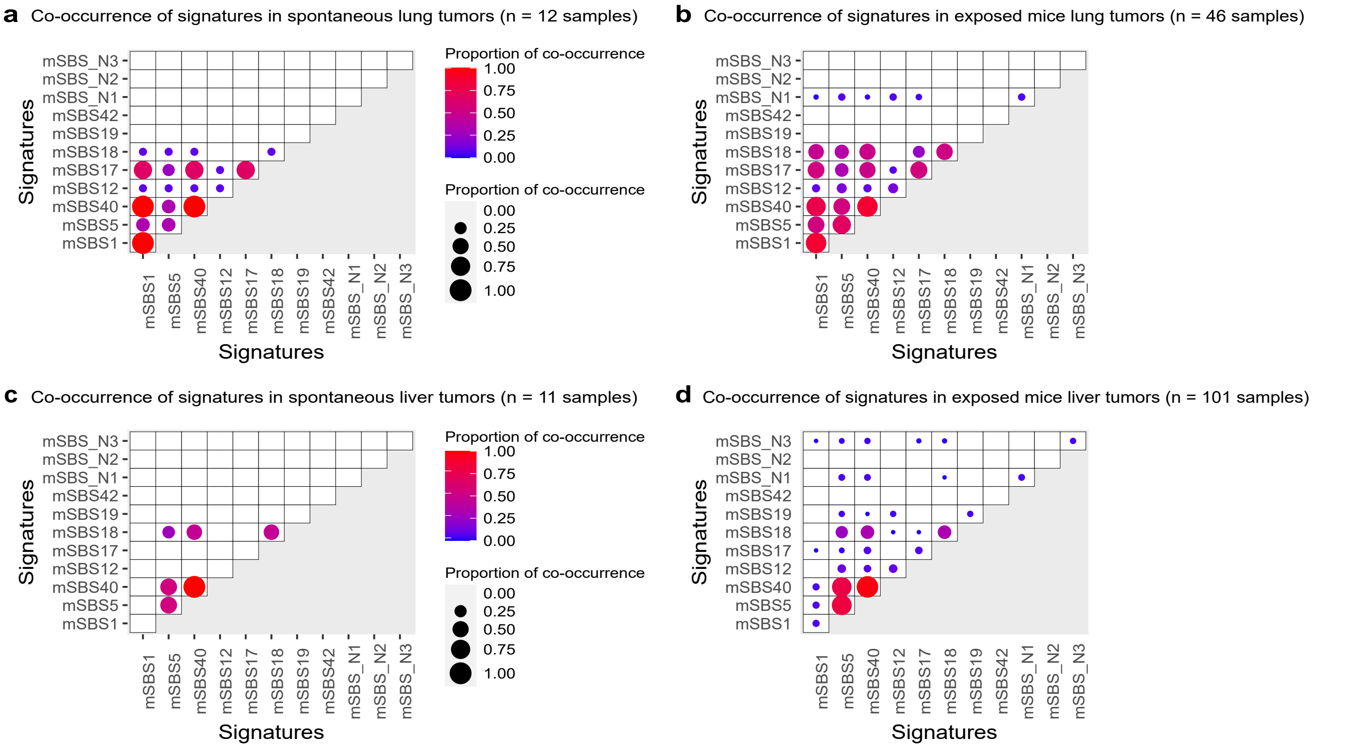


**Figure S2 | Effect of carcinogen exposure on the co-occurrence of mutational signatures, in mice exposed to 20 known or suspected carcinogens (data from[12]). a**, Co-occurrence of signatures in spontaneous lung tumors. **b**, Co-occurrence of signatures in lung tumors from exposed mice (all carcinogens considered). **c**, Co-occurrence of signatures in spontaneous liver tumors. **d**, Co-occurrence of signatures in liver tumors from exposed mice (all carcinogens considered).

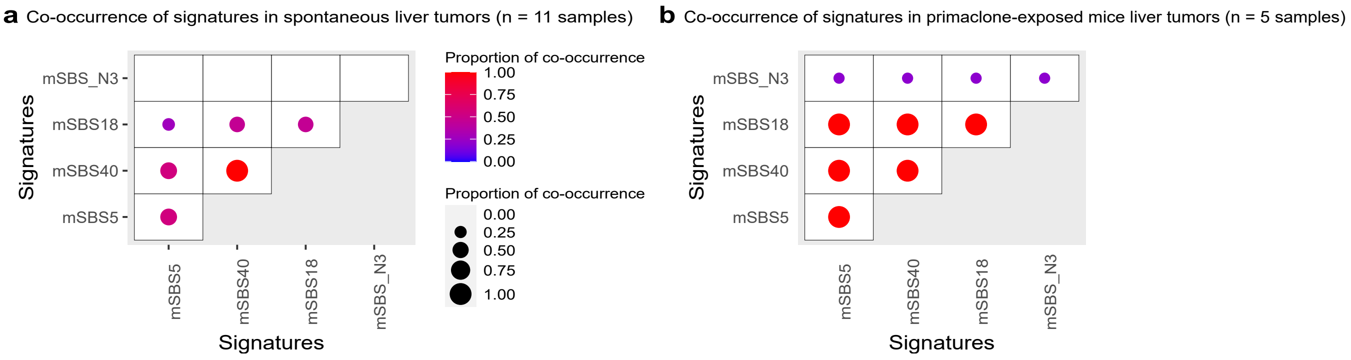


**Figure S3 | Effect of primaclone exposure on the co-occurrence of mutational signatures in mice (data from[12]). a**, Co-occurrence of signatures in spontaneous liver tumors. **b**, Co-occurrence of signatures in primaclone-exposed liver tumors.

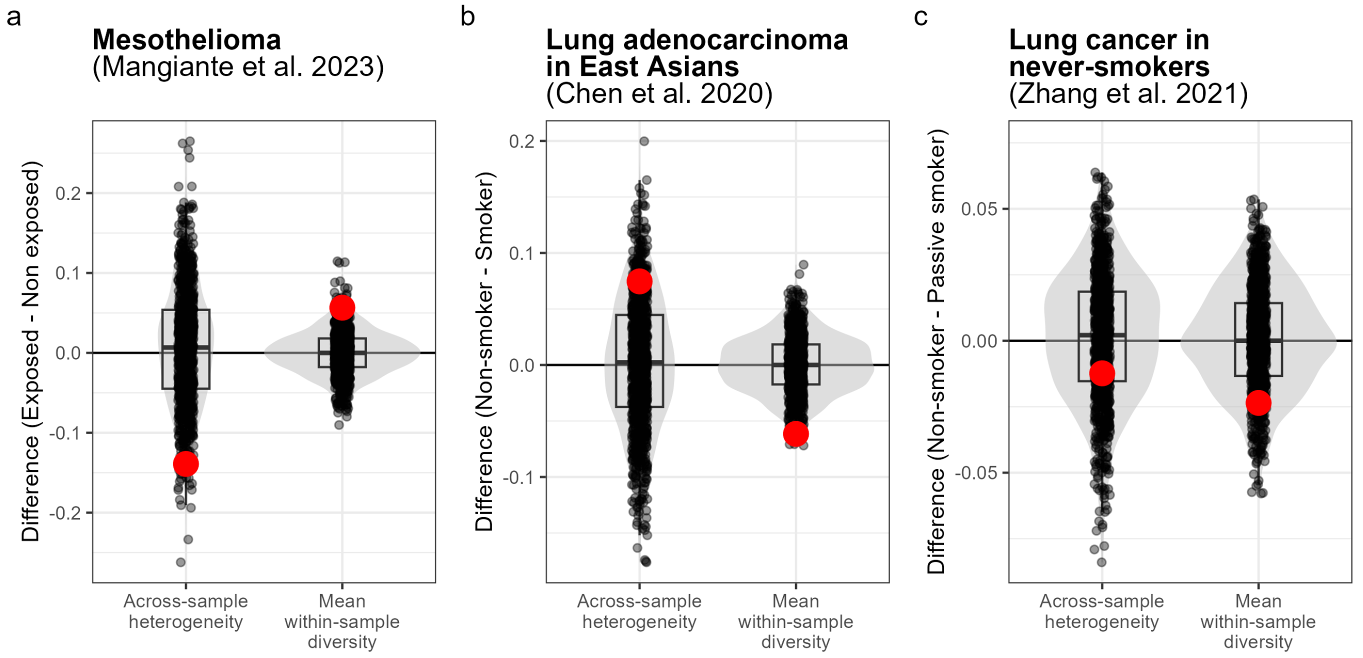


**Figure S4 | Bootstrap comparisons of exposure groups for the three datasets presented in Figure 2b.** In each panel, violin plots and box plots represent the null distributions of the differences between exposure groups in either across-sample heterogeneity (left) or mean within-sample diversity (right). These distributions came from drawing bootstrap replicates (black dots) of each exposure group under the null hypothesis that there is no difference between groups. Bootstrapping is explained in the methods section. Red dots represent the true difference between exposure groups observed in the data. When the true difference is negative, the one-sided $P$-value is computed as the fraction of bootstrap replicate differences less than the true difference; when the true difference is positive, the one-sided $P$-value is the fraction of replicate differences that exceed the true difference. **a,** Bootstrap distributions of signature variability differences between mesothelioma samples exposed or non-exposed to asbestos.[14] 22/1,000 bootstrap replicates have a smaller difference in across-sample variability, and 28/1000 have a greater difference in mean within-sample diversity than the true differences, leading to P-values of 0.022 and 0.028 for these one-sided comparisons. **b,** Bootstrap distributions of signature variability differences between lung adenocarcinoma samples in non-smokers and smokers.[24] 112/1,000 bootstrap replicates have a greater difference in across-sample variability, and 5/1000 have a lesser difference in mean within-sample diversity than the true differences, leading to P-values of 0.112 and 0.005 for these one-sided comparisons. **c,** Bootstrap distributions of signature variability differences between lung cancer samples in non-smokers or passive smokers.[11] 294/1,000 bootstrap replicates have a greater difference in across-sample variability, and 116/1000 have a lesser difference in mean within-sample diversity than the true differences, leading to P-values of 0.294 and 0.116 for these one-sided comparisons.

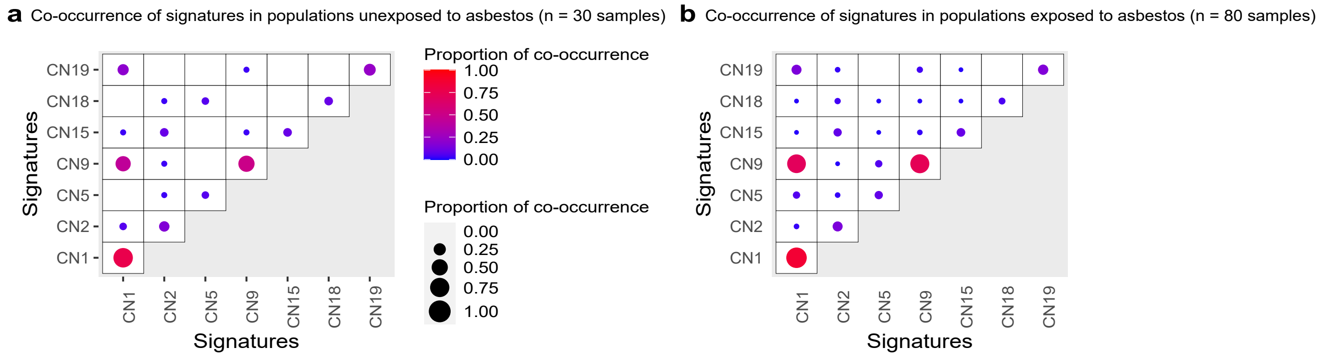


**Figure S5 | Effects of exposure to asbestos on the co-occurrence of mutational signatures in pleura (data from[14]). a**, Co-occurrence of signatures in non-exposed malignant pleural mesothelioma patients. **b**, Co-occurrence of signatures in exposed malignant pleural mesothelioma patients.

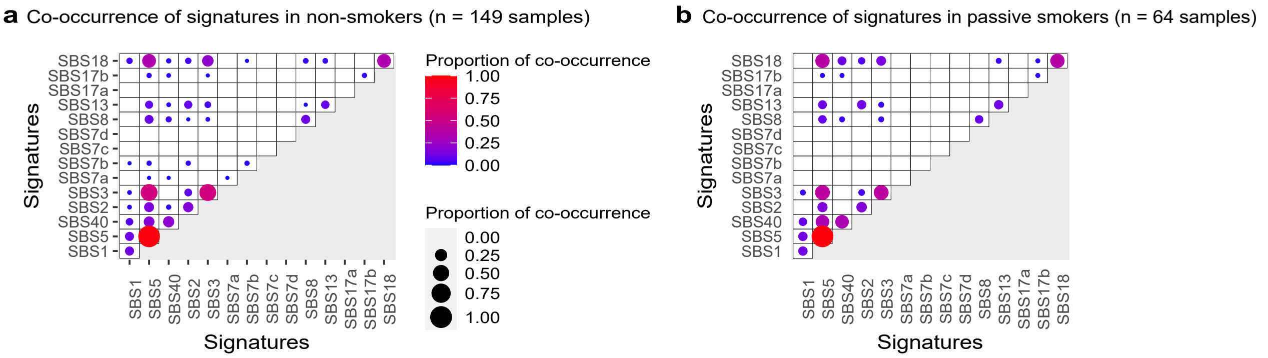


**Figure S6 | Effects of exposure to second-hand smoke on the co-occurrence of mutational signatures in lungs (data from[11]). a**, Co-occurrence of signatures in non-smoker lung cancer patients unexposed to second-hand smoke. **b**, Co-occurrence of signatures in non-smoker lung cancer patients exposed to second-hand smoke.

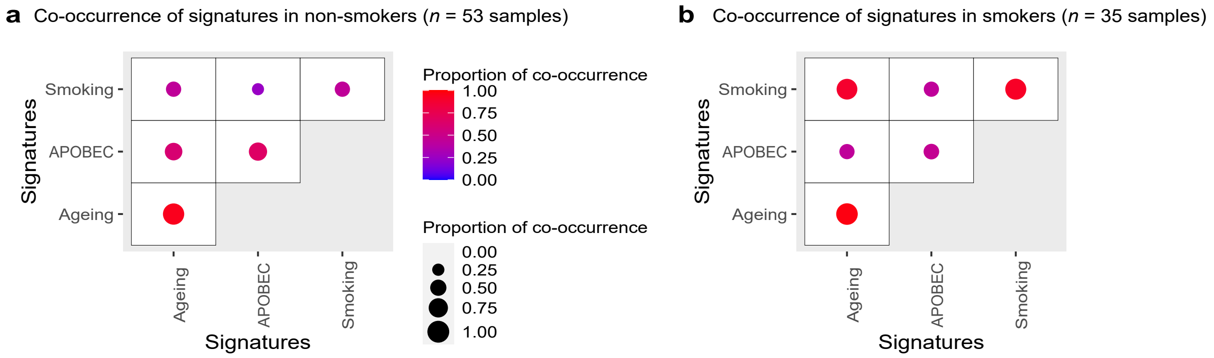


**Figure S7 | Effects of smoking on the co-occurrence of mutational signatures in lungs (data from[24]). a**, Co-occurrence of signatures in non-smoker East Asian patients with adenocarcinoma. **b**, Co-occurrence of signatures in smoker East Asian patients with adenocarcinoma.

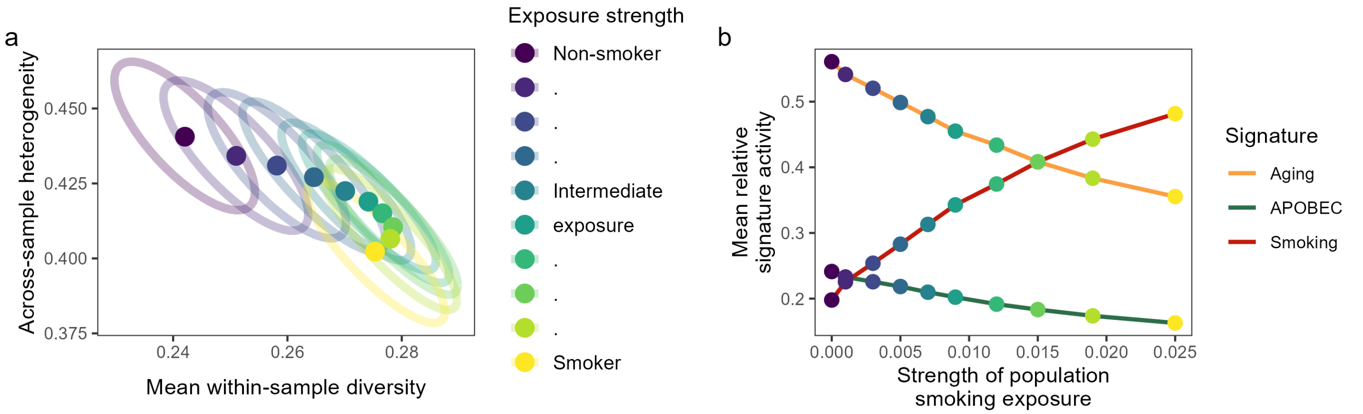


**Figure S8 | Results of simulations exploring the effect of smoking exposure on SVA results. a,** Relationship between mean population smoking exposure (color) and SVA results for simulated data. As the mean strength of exposure to the smoking mutational signature increases from the level of the non-smoking population of Chen and colleagues[24] to the level of the smoking population from the same study, a simulated population's mutational signature activity becomes more homogeneous across samples (one-sided t-test, $P<10^{-4}$ for all pairwise comparisons). The mean diversity of each individual's mutational signature activity increases until a threshold just before a smoker's exposure level, when it begins to decrease. However, all populations with more exposure than non-smokers have more within-sample signature diversity (one-sided t-test, $P<2\times10^{-16}$ for all comparisons). The ellipses represent the 90% confidence region of 500 simulations for each exposure strength. The simulation procedure is explained in the methods section. **b,** Relationship between the strength of population-level smoking exposure used in the simulation ($x$-axis, point color corresponds to color in panel a) and the mean relative activity of each mutational signature ($y$-axis). In panel a, mean within-sample diversity is maximized at a light-green exposure strength corresponding to strength $= 0.015$. We see in panel b that, when strength $= 0.015$, smoking overtakes aging as the dominant signature and the the mean abundance of the most abundant signature is minimized. When the dominant signature has relatively low abundance, the signature abundances are most even and diversity is maximized.

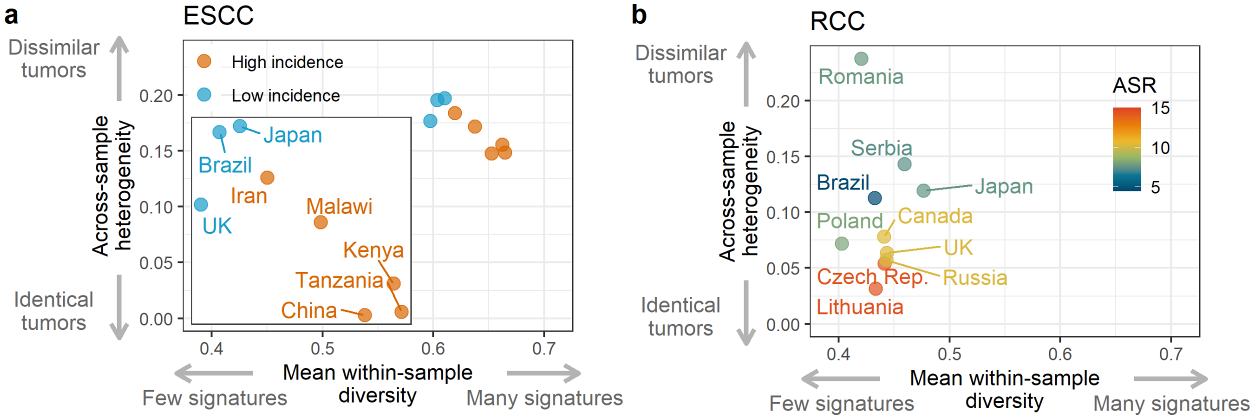


**Figure S9 | Signature variability analysis of esophageal squamous cell carcinoma (ESCC, a) and clear cell renal cell carcinoma (ccRCC, b), weighted by the cosine similarity among signatures.** ESCC data from[18]; ccRCC data from[25]. Analysis identical to that presented in Figure 3a,d, except that, here, both variability statistics account for the pairwise cosine similarity among mutational signatures. Novel SBS signatures with a 5-nucleotide context are reduced to their 3-nucleotide analog in order to compare to COSMIC signatures, leading SBS40a, b, and c to have high cosine similarities.

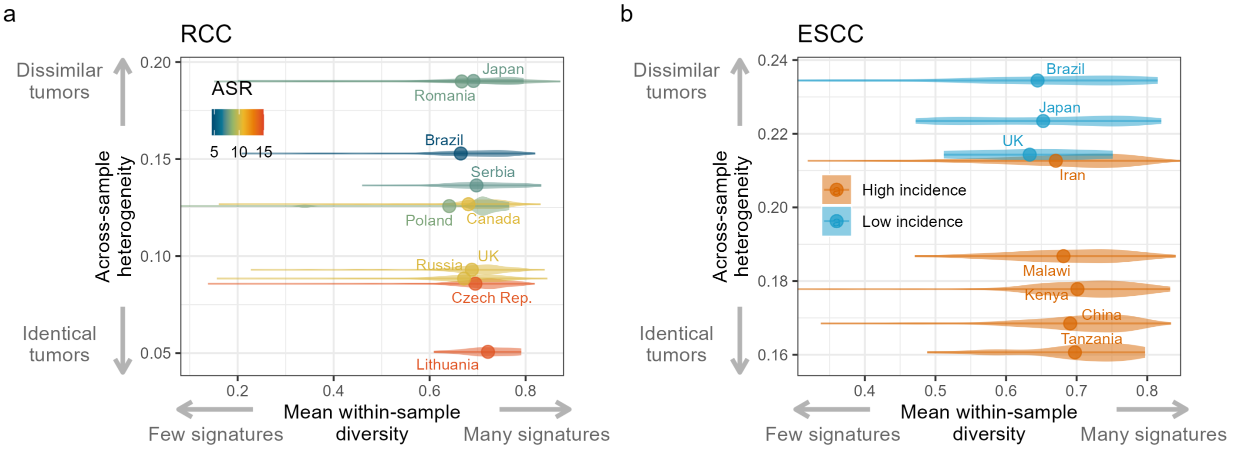


**Figure S10 | Signature variability analysis of esophageal squamous cell carcinoma (ESCC, a) and clear cell renal cell carcinoma (ccRCC, b), with violin plots showing the distributions of within-sample diversity values.** ESCC data from[18]; ccRCC data from[25]. Analysis identical to that presented in Figure 3b,e. For clarity, the x-axis is truncated to focus on the middle 95% of all distributions.

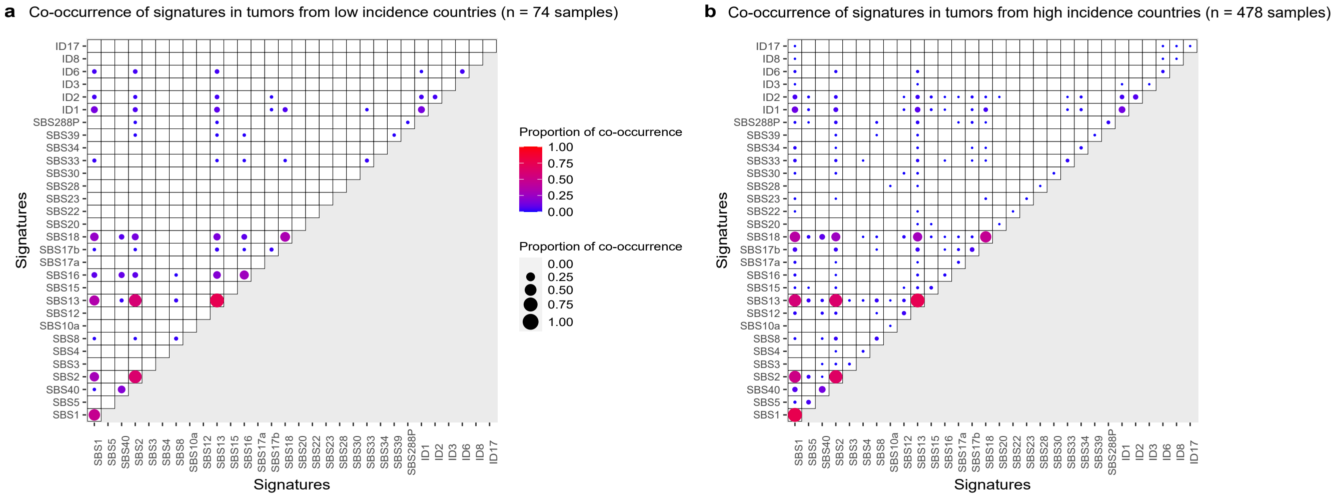


**Figure S11 | Effects of the regional level of esophageal squamous cell carcinoma incidence on the co-occurrence of mutational signatures in patients (data from[18]). a**, Co-occurrence of signatures in ESCC tumors from low incidence countries. **b**, Co-occurrence of signatures in ESCC tumors from high incidence countries.

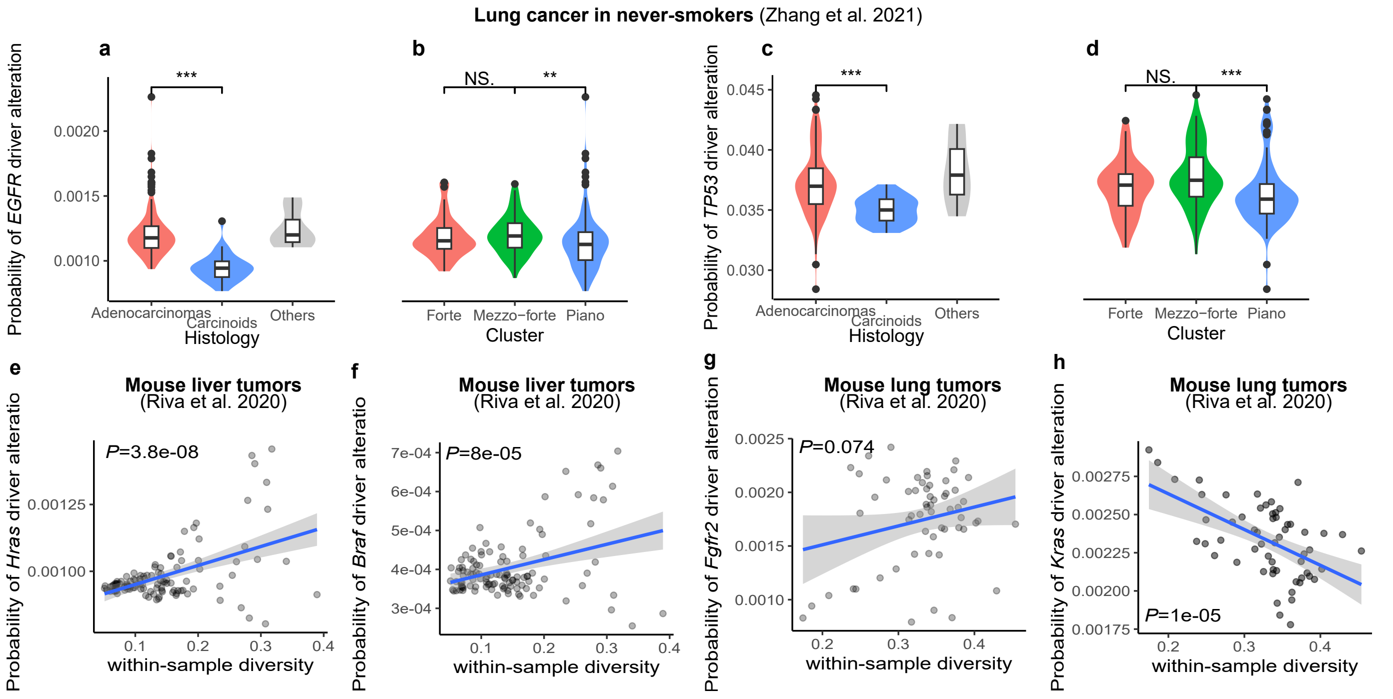


**Figure S12 | Within-sample diversity impacts driver mutations. a**-**b**, associations histology, copy number cluster, and the probability of a driver alteration in *EGFR*. Stars indicate the significance level of two-sided Wilcoxon rank sum tests: $*$ for $P<0.05$, $**$ for $P<0.01$, and $***$ for $P<0.001$. In box plots, the center line represents the median, box limits represent the upper and lower quartiles, whiskers represent 1.5 times the interquartile range, and points represent outliers. **c**-**d**, Same as a-b but for gene *TP53*. **e**-**h**, Association between within-sample diversity and the probability of a driver alteration in each of the four cancer genes with recurrent hotspots mutations identified by Riva and colleagues[12].

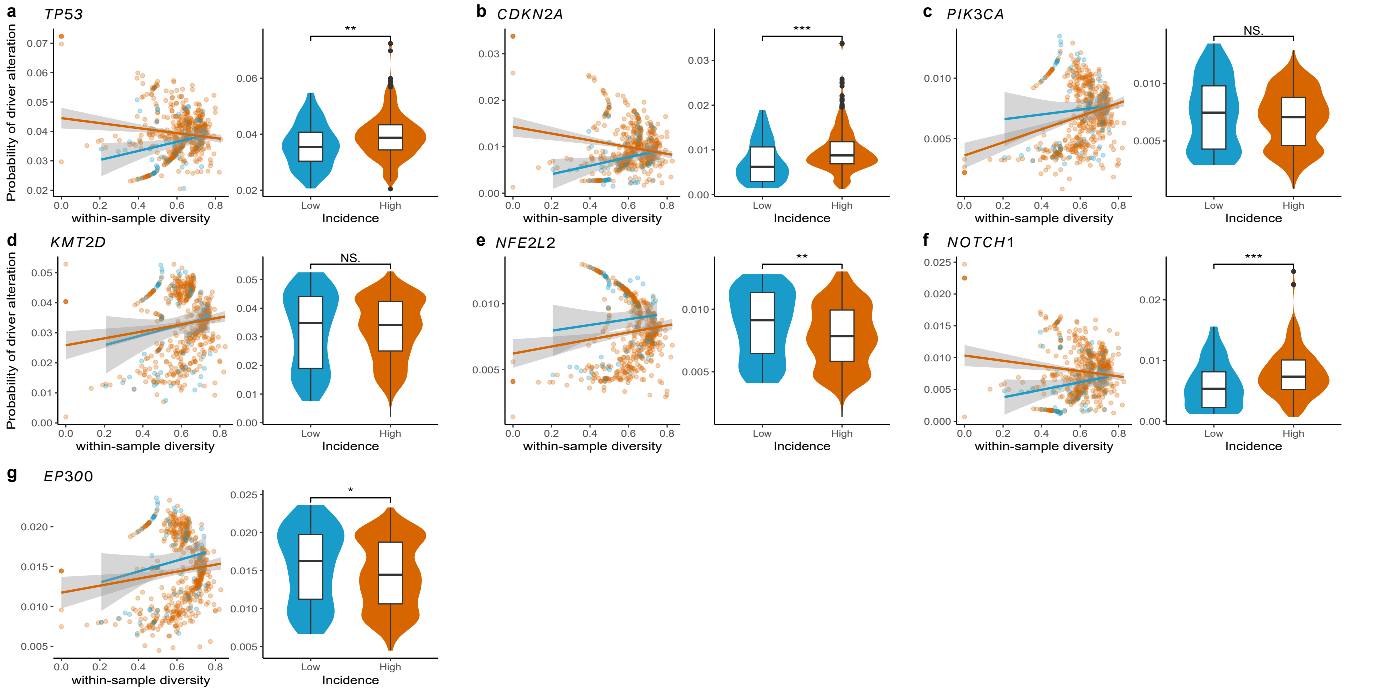


**Figure S13 | Within-sample diversity impacts driver mutation probability in ESCC across countries with varying incidence. a**, Association between within-sample diversity and the probability of a mutation being a driver mutation in gene *TP53* in ESCC across countries with varying incidence (data from[18]). Stars indicate the significance level of two-sided Wilcoxon rank sum tests: NS for $P > 0.05$, $*$ for $P<0.05$, $**$ for $P<0.01$, and $***$ for $P<0.001$. **b**-**g**, Same as a but for genes *CDKN2A*, *PIK3CA*, *KMT2D*, *EP300*, *NFE2L2*, and *NOTCH1*. In box plots, the center line represents the median, box limits represent the upper and lower quartiles, whiskers represent 1.5 times the interquartile range, and points represent outliers.

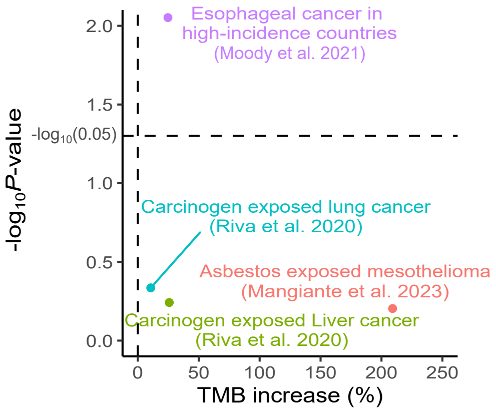


**Figure S14 | Analysis of tumor mutational burden (TMB) across datasets.** Increase in TMB is measured as the mean TMB difference between tumors in a reference group and tumors in the alternative group (positive when TMB is greater in samples from the alternative group, negative otherwise), expressed as a percentage of the TMB in the reference group. $P$-values correspond to two-sided $t$-tests comparing the TMB in the two groups.For Esophageal cancers, the reference are tumors from low-incidence countries and the alternative group is tumors from high-incidence countries. For Other datasets, the reference is unexposed individuals and the alternative group is exposed individuals.

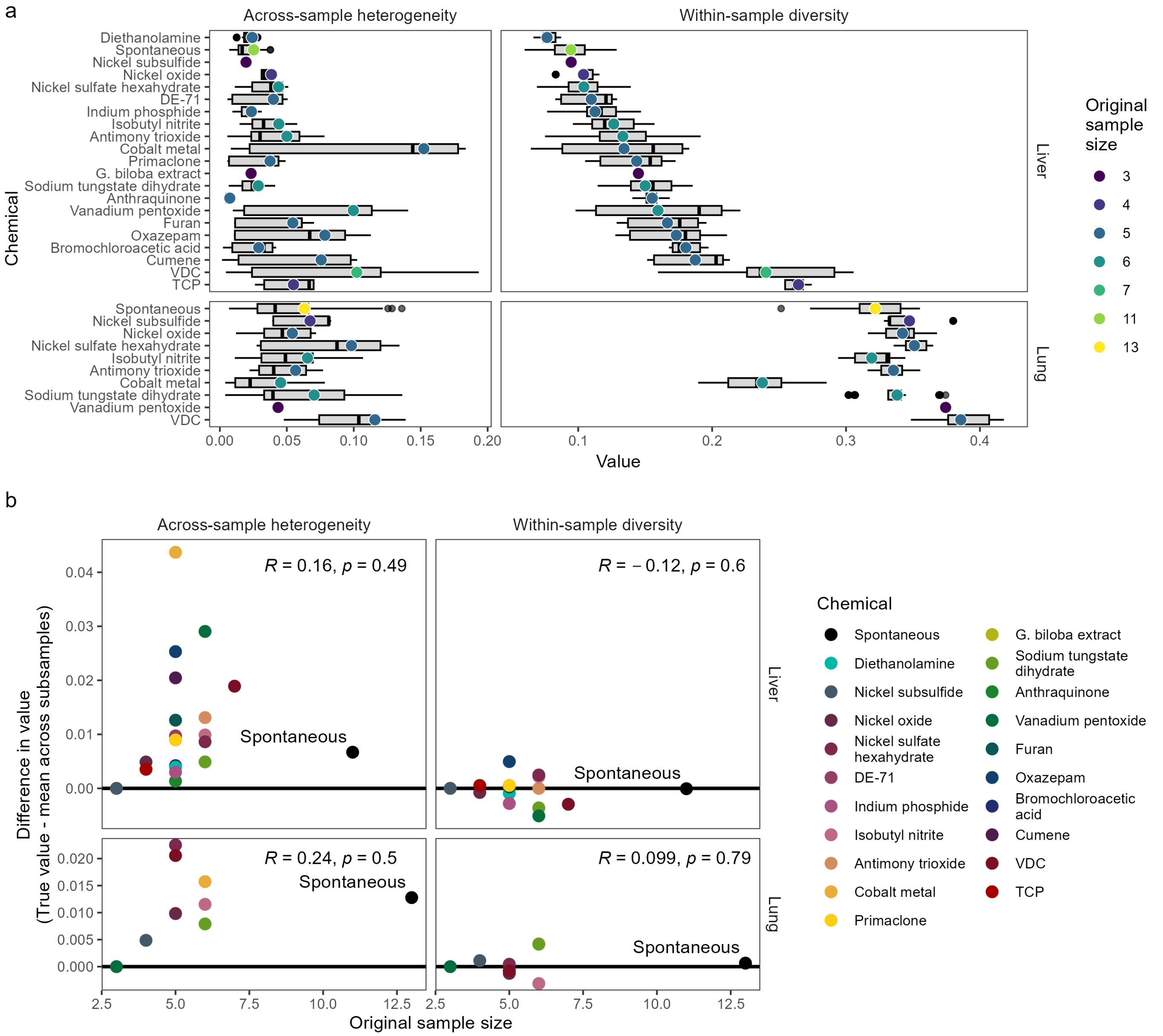


**Figure S15** | **SVA differences between spontaneous and carcinogen-induced tumors are not biased by sample size. a,** Points denote the value of across-sample heterogeneity (left) and within-sample diversity (right) for mouse tumors in the liver (top) or lung (bottom), caused by each chemical ($y$-axis). Point color represents the original sample size for each chemical and organ combination, also presented in table S2. Box plots convey the distribution of SVA results when each data set is subsampled to three samples (without replacement) 500 times. The center line represents the median, box limits represent the upper and lower quartiles, whiskers represent 1.5 times the interquartile range, and points represent outliers. **b,** Relationship between original number of tumors sampled for each exposure group and the difference between the true variability of each exposure group (i.e., each point in panel a) and the mean variability of that exposure group when subsampled to 3 tumors (i.e., the mean of each distribution in panel a). These differences are centered at approximately 0 for within-sample diversity, but are centered at greater than 0 for across-sample heterogeneity, suggesting that, across chemical types and sample sizes, heterogeneity is greater when the sample size exceeds 3. Although spontaneous tumors are in outlier in their sample size, they do not stand out in their susceptibility to sampling bias.

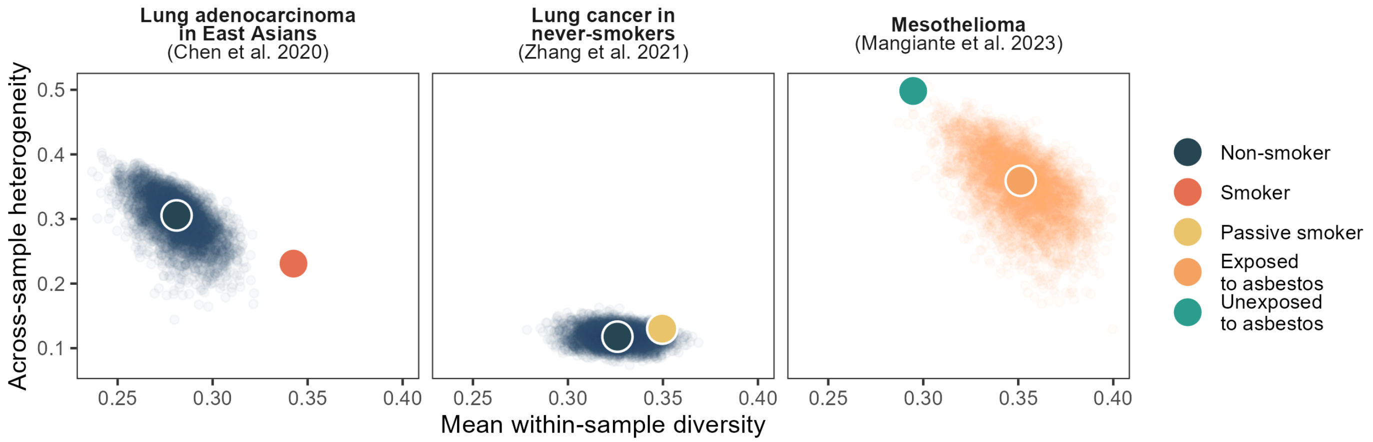


**Figure S16 | Comparisons of exposure groups are not biased by differences in sample size.** The exposure groups in each of the three data sets presented in Figure 2b differ in sample sized: lung adenocarcinoma in East Asians[24] (53 non-smokers, 35 smokers), LCINS[11] (149 non-smokers, 64 passive smokers), and mesothelioma[14] (80 exposed to asbestos, 30 unexposed to asbestos). In order to explore the effect of these differences in sample size, we subsampled the larger exposure group down to the size of the smaller exposure group for each data set, and conducted SVA on the smaller subset. Clouds of points give 5,000 subsampled replicates for each exposure group. Large points give SVA results for the full data. In all cases with significant differences in SVA results between exposure groups (lung adenocarcinoma in East Asians and mesothelioma), the true SVA values are near the center of the cloud of subsampled points, while the opposite exposure group is not contained within the point cloud, suggesting that the difference between exposure groups is not driven by sampling differences between exposure groups.

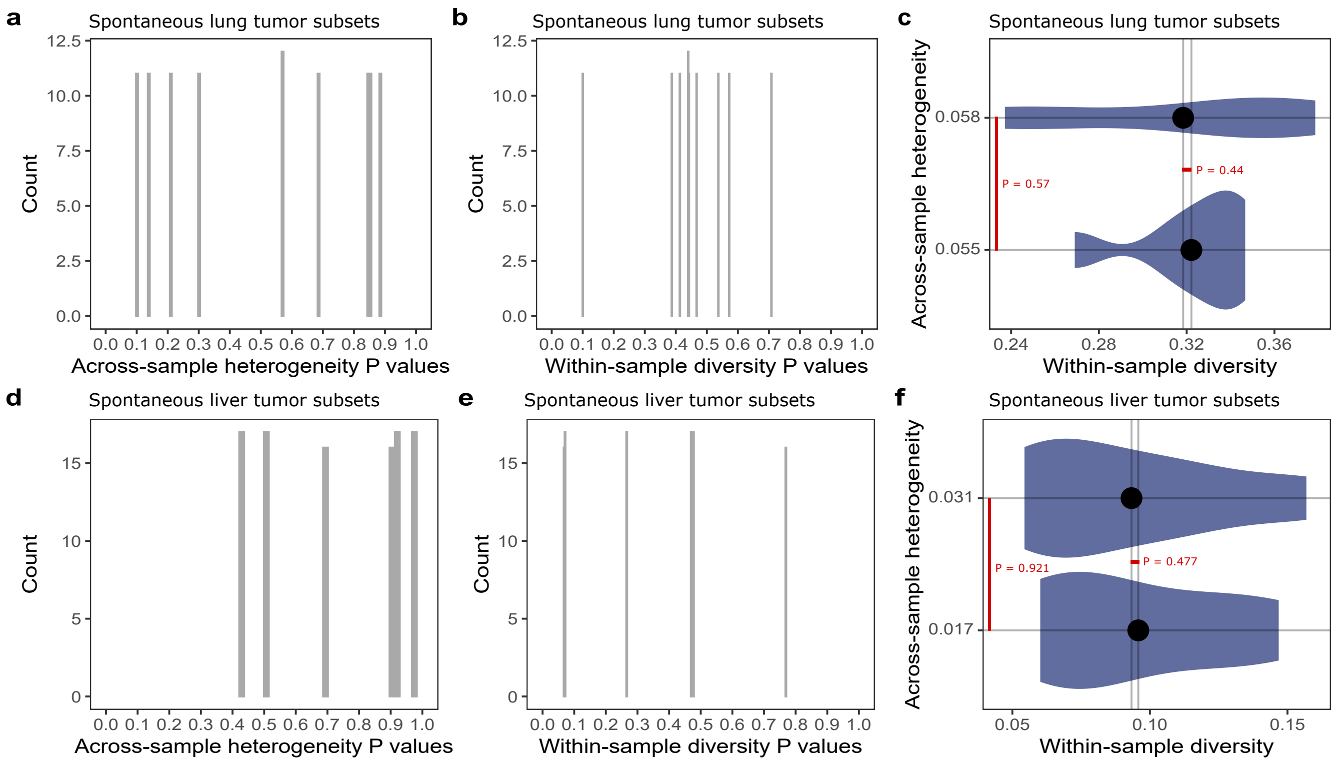


**Figure S17 | Null test of SVA using spontaneous mouse tumors (data from[12]).** **a**, Distribution of across-sample heterogeneity one-sided $P$ values after 100 repetitions of SVA comparing two random subsets of the 12 spontaneous mouse lung tumor samples using bootstrapping (see methods). **b**, Distribution of within-sample diversity $P$ values after 100 repetitions of SVA comparing two subsets of all 12 spontaneous mouse lung tumor samples. **c**, Violin plot illustrating one of the 100 analyses performed for the null test. The spontaneous mouse lung tumor samples were split in 2 subsets (6 samples each) and compared using SVA. **d**-**f** Same as a-c but using the 12 spontaneous liver tumors.

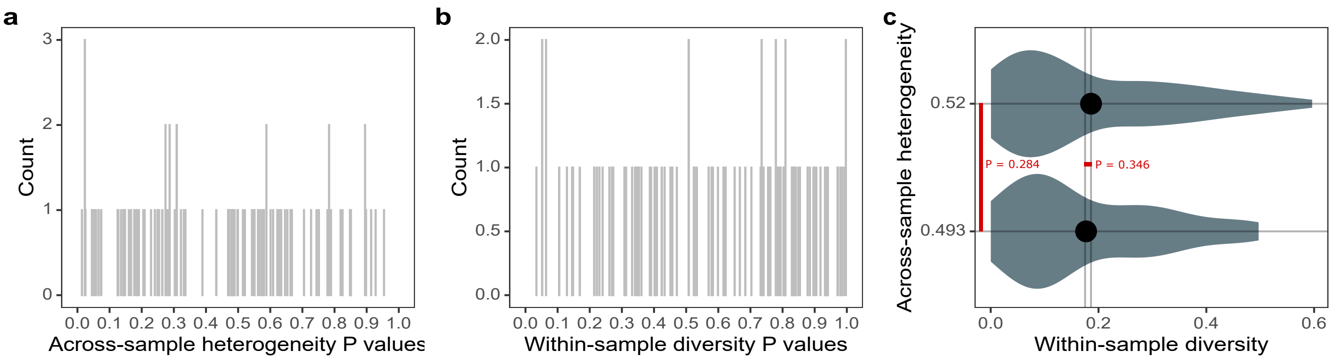


**Figure S18 | Null test of SVA using samples simulated under a Dirichlet distribution fitted on never-smoker samples unexposed to second-hand smoke (data from[11]; see methods). a**, Distribution of across-sample heterogeneity two-sided $P$ values after 100 repetitions of SVA comparing two subsets of simulated tumor samples. **b**, Distribution of within-sample diversity two-sided $P$ values after 100 repetitions of SVA comparing two subsets of simulated tumor samples. **c**, Violin plot illustrating one of the 100 analyses performed for the null test. The simulated samples were split in 2 subsets and compared using SVA.

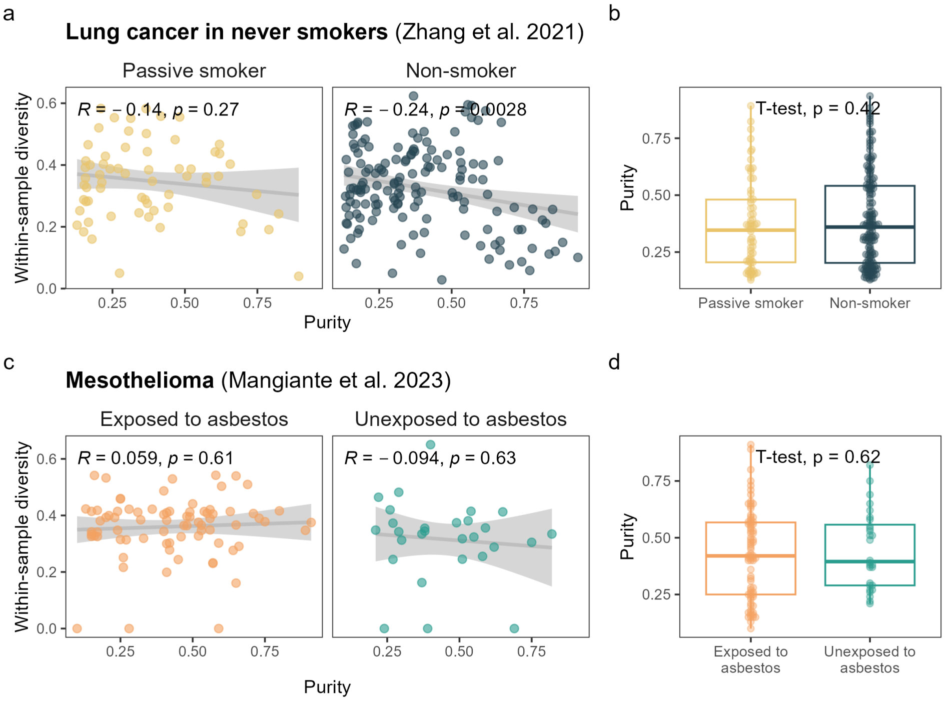


**Figure S19 | Purity does not drive differences in SVA results between high- and low-carcinogen-exposure populations. a,** Association between within-sample signature diversity and purity in never smokers exposed to second-hand smoke ("Passive smoker") and in unexposed never smokers ("Non-smoker"). **b,** Purity in samples from passive smokers and non-smokers. **c,** Association between within-sample signature diversity and purity in mesothelioma samples from people exposed or unexposed to asbestos. **d,** Purity in mesothelioma samples from asbestos-exposed and unexposed people. In a and c, $P$ values correspond to two-sided Pearson correlation tests and the gray area corresponds to 95% confidence bands on the slope and intercept of an ordinary least-squares regression line. In b and d, $P$ values corresponds to a Welch two-sample two-sided t-test. In box plots, the center line represents the median, box limits represent the upper and lower quartiles, whiskers represent 1.5 times the interquartile range, and points represent outliers. Data presented reflects two of the examples presented in Figure 2b.

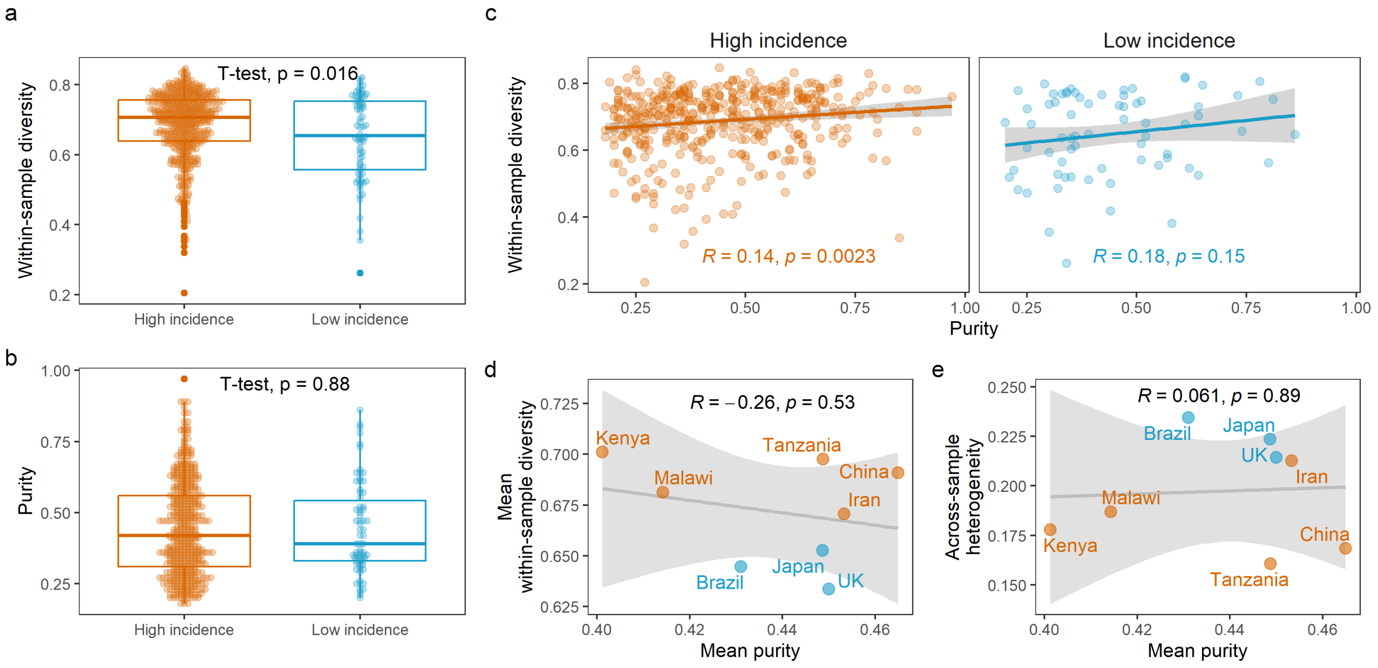


**Figure S20 | Purity does not drive differences in SVA results between high- and low-ESCC-incidence countries. a,** Within-sample signature diversity in samples from high- and low-incidence countries. **b,** Purity in samples from high- and low-incidence countries. In a and b, $P$ values corresponds to a Welch two-sample two-sided t-test. In box plots, the center line represents the median, box limits represent the upper and lower quartiles, whiskers represent 1.5 times the interquartile range, and points represent outliers. **c,** Association between within-sample signature diversity and purity. **d,** Association between the mean purity of samples in a country and the mean within-sample signature diversity of those samples. **e,** Association between the mean purity of samples in a country and the across-sample signature heterogeneity of those samples. In d and e, $P$ values correspond to two-sided Pearson correlation tests. In c–e, the gray area corresponds to 95% confidence bands.

**Table S1 | One-sided bootstrap P-values comparing SVA results for each tissue and chemical to those of spontaneous tumors of the same tissue.**

| | Across-sample heterogeneity | | Mean within-sample diversity | |
|---|---|---|---|---|
| **Chemical** | **Liver** | **Lung** | **Liver** | **Lung** |
| Anthraquinone | 0.873 | - | ***0.001*** | - |
| Antimony trioxide | 0.142 | 0.46 | ***0.072*** | 0.271 |
| Bromochloroacetic acid | 0.369 | - | ***0.001*** | - |
| Cobalt metal | ***0.066*** | 0.592 | 0.152 | 1 |
| Cumene | 0.156 | - | ***0.01*** | - |
| DE-71 | 0.144 | - | 0.234 | - |
| Diethanolamine | 0.396 | - | 0.85 | - |
| Furan | 0.178 | - | ***0.013*** | - |
| *G. biloba* extract | 0.288 | - | ***0.032*** | - |
| Indium phosphide | 0.409 | - | 0.213 | - |
| Isobutyl nitrite | ***0.072*** | 0.414 | ***0.05*** | 0.523 |
| Nickel oxide | 0.164 | 0.5 | 0.324 | 0.168 |
| Nickel subsulfide | 0.405 | 0.286 | 0.479 | 0.162 |
| Nickel sulfate hexahydrate | ***0.026*** | 0.166 | 0.312 | ***0.072*** |
| Oxazepam | 0.141 | - | ***0.019*** | - |
| Primaclone | 0.187 | - | ***0.045*** | - |
| Sodium tungstate dihydrate | 0.342 | 0.33 | ***0.012*** | 0.237 |
| TCP | 0.279 | - | ***<0.001*** | - |
| Vanadium pentoxide | 0.139 | 0.478 | ***0.057*** | ***0.022*** |
| VDC | ***0.064*** | 0.113 | ***0.001*** | ***0.007*** |
| **# of chemicals per tissue:** | 20 | 9 | 20 | 9 |
| **# of significant chemicals ($P<0.1$):** | 4 | 0 | 13 | 3 |
| **# of significant chemicals ($P<0.05$):** | 1 | 0 | 10 | 2 |

The alternative hypothesis is that spontaneous tumors are *less* variable than exposed tumors. Low P-values for a chemical, tissue (liver or lung), and variability type (across-sample or mean within-sample) suggest that, for that chemical and tissue combination, we can reject the null hypothesis of there being no difference in that type of variability between spontaneous and exposed tumors, concluding that there is less variability in spontaneous tumors than in exposed tumors. Note that there is one case where $P > 0.9$ (Cobalt metal, lung, mean within-sample diversity), suggesting that spontaneous lung tumors have significantly *more* within-sample diversity than do lung tumors caused by cobalt metal.

**Table S2 | Number of mouse tumor samples in each exposure group presented in Figure 2a.**

| Chemical | Liver | Lung |
|---|---|---|
| Spontaneous | 11 | 12 |
| Anthraquinone | 5 | - |
| Antimony trioxide | 6 | 5 |
| Bromochloroacetic acid | 5 | - |
| Cobalt metal | 5 | 6 |
| Cumene | 5 | - |
| DE-71 | 5 | - |
| Diethanolamine | 5 | - |
| Furan | 5 | - |
| *G. biloba* extract | 3 | - |
| Indium phosphide | 5 | - |
| Isobutyl nitrite | 6 | 6 |
| Nickel oxide | 4 | 5 |
| Nickel subsulfide | 3 | 4 |
| Nickel sulfate hexahydrate | 6 | 6 |
| Oxazepam | 5 | - |
| Primaclone | 5 | - |
| Sodium tungstate dihydrate | 6 | 6 |
| TCP | 4 | - |
| Vanadium pentoxide | 6 | 3 |
| VDC | 7 | 5 |

Data from[12].

**Table S3 | Co-occurrence of signatures.** Available in Supplementary_tables.xlsx

**Table S4 | Associations between signature diversity, exposure status, and covariates for carcinogen exposure data sets.** Available in Supplementary_tables.xlsx

**Table S5 | Tests of association between incidence and SVA results for ESCC and RCC.** Available in Supplementary_tables.xlsx

**Table S6 | Linear regression between country-level mean mutational signature abundances and SVA results for ESCC.** Available in Supplementary_tables.xlsx

**Table S7 | Linear regression between country-level mean mutational signature abundances and SVA results for RCC.** Available in Supplementary_tables.xlsx

**Table S8 | Probability of driver mutations in LCINS.** Available in Supplementary_tables.xlsx

**Table S9 | Associations between probability of driver alterations, signature diversity, exposure status, and covariates for LCINS.** Available in Supplementary_tables.xlsx

**Table S10 | Probability of driver mutations in mice exposed to known and suspected carcinogens.** Available in Supplementary_tables.xlsx

**Table S11 | Probability of driver mutations in ESCC across continents.** Available in Supplementary_tables.xlsx

**Table S12 | Associations between probability of driver alterations, signature diversity and incidence group for ESCC across countries** Available in Supplementary_tables.xlsx

**Table S13 | Association between carcinogen exposure and TMB.** Available in Supplementary_tables.xlsx
